# Adolescent health and not in education, employment or training (NEET) in young adulthood: Evidence from a UK prospective longitudinal study

**DOI:** 10.64898/2026.08.13.26360381

**Authors:** Dominic P. Kelly, Jacques Wels, Praveetha Patalay

## Abstract

**Background:** High rates of young people who are not in education, employment or training (NEET) are a major societal concern in the UK. Whilst other studies have highlighted that adolescent health can predict NEET status in young adulthood, robust and recent longitudinal evidence remains limited.

**Methods:** This study used data from the Millennium Cohort Study, a longitudinal study of people born in the UK in the early 2000s, to examine associations between adolescent mental health difficulties and childhood neurodevelopmental conditions, indicators of physical health and health behaviours and NEET status in early adulthood (median age: 23). NEET status was derived from participant reports of economic and education activity. Co-occurrence of exposures was also considered, and population attributable fractions were calculated to account for differences in exposure prevalence.

**Results:** Among 8,135 young people, 12.1% were NEET at age 23. Estimates adjusted for demographic factors indicated that multiple health exposures increased risk of being NEET at age 23, with larger associations observed with most mental health difficulties and childhood neurodevelopmental conditions compared to physical health and health behaviour exposures. For instance, a longstanding mental health condition more than doubled the risk of being NEET (adjusted relative risk [aRR] = 2.15, 95% CI = 1.74, 2.65), while autism (aRR = 2.94, 95% CI = 2.31, 3.74) and ADHD (aRR = 2.74, 95% CI = 2.11, 3.56) almost tripled the risk. Reports of greater number of mental health and neurodevelopmental difficulties by adolescence were associated with greater risk of being NEET in young adulthood. Obesity predicted being NEET at age 23 (aRR = 1.48, 95% CI = 1.20, 1.83) and obesity accompanied by a mental health or neurodevelopmental difficulty further increased risk (aRR = 1.95, 95% CI = 1.47, 2.58).

**Conclusions:** Findings indicate that health exposures by adolescence, especially mental and neurodevelopmental difficulties, are strongly associated with being NEET in early adulthood.

## Background

As of 2025, approximately one in eight young adults in the UK, typically defined as those aged 18 to 25, are not in education, employment or training (NEET; 1). Young adults who are NEET report poorer health, on average, than their peers (2) but this association is complex and bidirectional (3). Poorer health has been associated with a greater risk of subsequent NEET status (4). Therefore, determining key adolescent health exposures may help identify young people who would benefit most from earlier intervention efforts to reduce the risk of later NEET status. Longitudinal datasets, especially birth cohorts, are vital resources for determining childhood and adolescence predictors of adult outcomes.

Gen Z (the generation born between 1997 and 2012) entered a changing labour market affected by the development of the gig economy, the use of zero-hours contracts and, more recently, a reconfiguration due to the widening use of artificial intelligence. Gen Z also experienced unique social and economic circumstances and has a higher reported prevalence of mental health conditions and neurodevelopmental conditions such as attention deficit hyperactivity disorder (ADHD) and autism than previous generations (5, 6). Economic changes do not affect all young people equally and young people who are already disadvantaged may be disproportionately affected.

Meta-analytic evidence indicates that young people with mental health difficulties have 28% higher odds of being NEET than those without (5). However, mental health is multidimensional and pooling different mental health conditions into single estimates may obscure important differences. Existing longitudinal evidence suggests that earlier externalising problems, such as conduct problems (6), and internalising problems, such as suicidal self-harm (7), each predict later NEET status.

There are few existing studies on the impact of a childhood autism diagnosis on being NEET, but evidence does point to autistic children having lower educational achievement and engagement, which is known to be linked to an increased likelihood of NEET status (8). Evidence also suggests that children with ADHD have increased risk for being NEET as adults (9).

Labour market entry may be particularly challenging for young people with physical health problems, yet comparatively less research has examined associations between NEET status and physical health conditions (see 10 for a systematic review of the limited evidence). Adolescent obesity has been associated with depression and lower self-esteem (11) and these effects may continue, or have consequences, for education and employment participation in adulthood. Examining the potentially additive effects of obesity with a mental health and neurodevelopmental difficulty allows for a better understanding of how physical and mental health interact. Adolescent physical inactivity could also serve as a proxy for physical health problems.

Adolescent engagement with addictive and risky health behaviours may also predict later NEET status. Longitudinal evidence suggests that adolescent cannabis use is associated with an increased risk of being NEET in young adulthood, but alcohol use is not (4). Research on other drugs typically treats substance use as a consequence rather than a predictor of NEET status, leaving the reverse association underexplored. Much of the existing evidence is based on cohorts before Gen Z and may not reflect health behaviours that are more relevant to contemporary adolescents. For example, although smoking has been linked to NEET status (12), declining smoking rates (13) means that vaping may be a more salient and underexplored predictor for Gen Z. Meta-analytic evidence also suggests an association between antisocial behaviour and NEET status (14).

In sum, relatively little is known about the health exposures during adolescence that predict economic activity during young adulthood for Gen Z specifically. This study addresses this knowledge gap by considering a wide range of health exposures in a contemporary UK cohort. The Millennium Cohort Study (MCS) is a nationally representative cohort study of children born in the UK between 2000 and 2002. The latest sweep of the MCS (15) provides an opportunity to examine whether data previously collected on different adolescent health exposures (16), both individually and in combination, help identify those at risk of being NEET in early adulthood (ages 21-24 years; henceforth referred to as age 23).

The aim of this study is to use the MCS data to better identify aspects of adolescent health that may help identify those suitable for targeted support to prevent later NEET status. We expect that mental health difficulties and childhood neurodevelopmental conditions will have stronger associations with later NEET status than indicators of physical health and health behaviours. Due to sex differences in the prevalence of health exposures (17) and trends in NEET over recent years, analyses were repeated sex-stratified to determine the degree to which associations between health and economic activity differ by sex.

## Methods

### Sample

The MCS sample was designed to overrepresent families of low socioeconomic status, families from minority ethnic backgrounds in England, and cohort members living in Scotland, Wales and Northern Ireland. Information was provided primarily by a parent during childhood and by the cohort members since age 17. For the first four sweeps, parents provided consent on the cohort members’ behalf; for all subsequent sweeps, the cohort member consented directly.

The original MCS sample consisted of 19,517 cohort members. Among these cohort members, 10,755 participated productively in the age 17 sweep and 8,489 of that group also participated productively in the age 23 sweep. Cohort members were excluded if they did not have valid data for NEET status at age 23 (n = 69), their data were unavailable (n = 47) or they did not provide information on exposures (n = 238), resulting in an analytic sample comprising 8,135 participants. As shown in *Supplementary Material A*, compared to participants who were not retained for the analytic sample, retained participants were more likely to be White, to be from a higher socioeconomic background and not to be NEET at age 17. Further details on participant non-response can be found in the technical reports for the age 17 and age 23 sweeps of the MCS (18, 19).

### Measures

#### Outcome

Cohort members reported their current economic activity at the time of interview during the age 23 sweep. Responses were classified into NEET or not NEET (see *Supplementary Material B* for coding scheme), in line with the UK Office for National Statistics’ definitions of employment, education and training (20).

#### Exposures

Health exposures were grouped into four domains: mental health difficulties, childhood neurodevelopmental conditions, indicators of physical health and health behaviours– see *Supplementary Material B* for more details.

Mental health difficulties assessed at age 17 included psychological distress (six-item Kessler Scale; 21) and indicators of conduct problems, emotional symptoms and hyperactivity (self-reported Strengths and Difficulties questionnaire; 22). Established cut-offs for high levels of difficulties were used. Self-harm in the past 12 months and details of having a longstanding mental health condition were also self-reported at age 17, as well as a suicide attempt by age 17.

**Figure 1.**
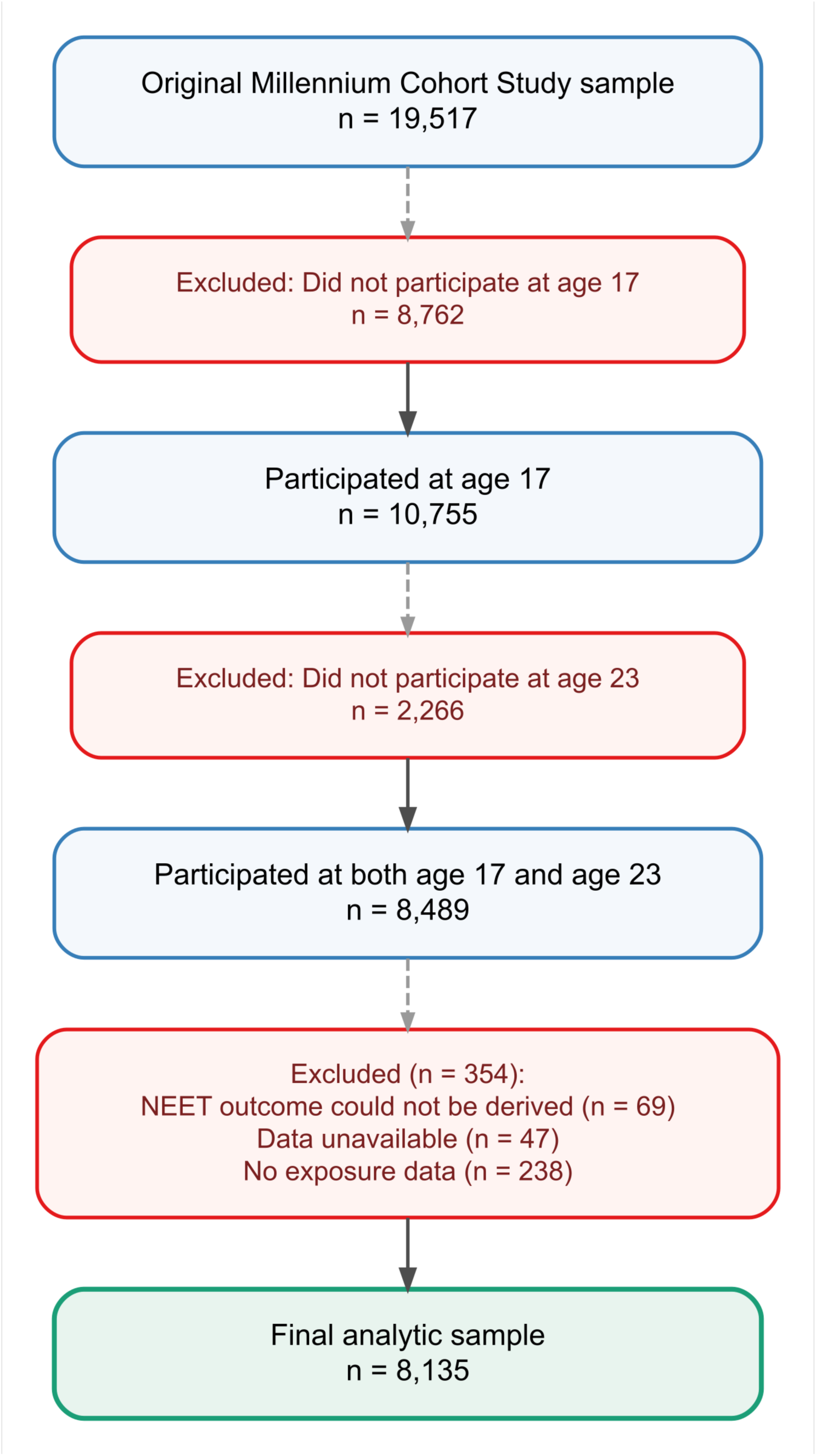
Flow chart showing the derivation of the analytic sample from the original Millennium Cohort Study sample.

For childhood neurodevelopmental conditions, autism and ADHD were assessed from parents’ reports drawn from data collection waves between ages 5 and 14 (23–26) of whether a health professional had informed them that their child had that condition.

Indicators of physical health included body mass index (BMI) scores categorised into not overweight, overweight and obese, using established cut-offs. Cohort members also self-reported physical health conditions and whether they reported at least one day in the previous week with one hour of moderate-to-vigorous physical activity.

All health behaviours, defined as modifiable lifestyle or risky behaviours, included were collected at age 17. Substance use was assessed by whether respondents used alcohol weekly, whether they binge drank repeatedly in the past year, whether they smoked or vaped at least occasionally, whether they had used cannabis in the past year and whether they had ever used hard drugs. Lastly, cohort members were asked to self-report whether they had participated in a list of antisocial behaviours in the past year. Other health behaviours of interest such as sleep and digital behaviours were considered but were only assessed in the web survey, which was completed by a smaller, selective subset of the participants. We therefore excluded these variables from the current analysis.

A composite measure of mental health and neurodevelopmental difficulties was constructed by summing for each cohort member indicators of high psychological distress, high conduct problems, a report of self-harm in the past 12 months, a suicide attempt by age 17, an autism diagnosis and an ADHD diagnosis. Responses were grouped into 0, 1, 2, or ≥3 difficulties. Three exposures were not included due to having strong theoretical overlap with other exposures in the composite: hyperactivity (overlap with ADHD), emotional symptoms (overlap with psychological distress) and a longstanding mental health condition (overlap with other symptoms and behaviours generally).

#### Covariates

Covariates included nation, sex, ethnicity, sexuality and four indicators of socioeconomic background from the cohort member’s childhood – highest parental occupation, highest parental qualification, whether the cohort member’s parents owned their home and whether their household income was below 60% of the OECD-equivalised (i.e., adjusting for household composition) median. NEET status was also collected at age 17 and used in a sensitivity analysis.

### Data Analysis

All analyses were conducted in R 4.3.2. All analyses were weighted using survey weights provided by the MCS to account for its complex sampling design and differential attrition. Descriptive statistics were used to estimate the prevalence of economic activities at age 23 and the prevalence of each demographic factor and adolescent health exposure. All p-values were corrected for multiple comparisons following the Holm procedure.

Relative risks (RR) with accompanying 95% confidence intervals (95% CIs) were estimated using Poisson regression models with robust standard errors to examine associations between each adolescent health exposure and NEET status at age 23. Results are reported before and after adjustment for demographic factors.

Because the prevalence of adolescent health exposures varied substantially, population attributable fractions (PAF) were also estimated using adjusted RRs to illustrate population-level patterns in associations. These were calculated using Miettinen’s formula *PAF = p_c_* (1-1/*RR*), where *p_c_* is the weighted prevalence of exposure among NEET cases (27). PAFs were only calculated for associations that increased the risk of NEET status at age 23 and had a significant p-value after correcting by the Holm procedure.

Secondary analyses stratified the sample by sex and NEET subtype and repeated the same models as the primary analyses. Adjusted relative risk analyses stratified by sex were followed up by including an interaction term between sex and each exposure. Joint Wald tests were used to determine if the interactions were statistically significant (*p* < .05).

We addressed missing data in the exposures and covariates using multiple imputation by chained equation (MICE) with the *mice* package in R (28). Missing data were limited – all health exposures had less than 10% missing data and among the covariates, only parents’ occupation exceeded 10% (18.2%). Thirty imputed datasets were generated; analyses were conducted separately and then pooled together using Rubin’s rules. The outcome was not imputed. Auxiliary variables included prior indicators of mental health, physical health, substance use and participation in prior waves. Composite measures were not imputed but were reconstructed within each imputed dataset after imputation. PAFs were calculated by combining the adjusted relative risks from the imputed models with the non-imputed weighted exposure prevalence among NEET cases.

## Results

*Table 1* presents the demographics of the analytic sample before and after applying population weights. 12.1% (95% CI: 11.0%, 13.2%) of the weighted sample of cohort members were NEET at age 23, including 7.9% who were unemployed and seeking work (95% CI: 7.2%, 8.6%) and 4.2% (95% CI: 3.5%, 4.9%) who were economically inactive. Among the remainder, 72.1% (95% CI: 70.6%, 73.7%) were employed and 15.8% (95% CI: 14.6%, 16.9%) were in education.

**Table 1.**
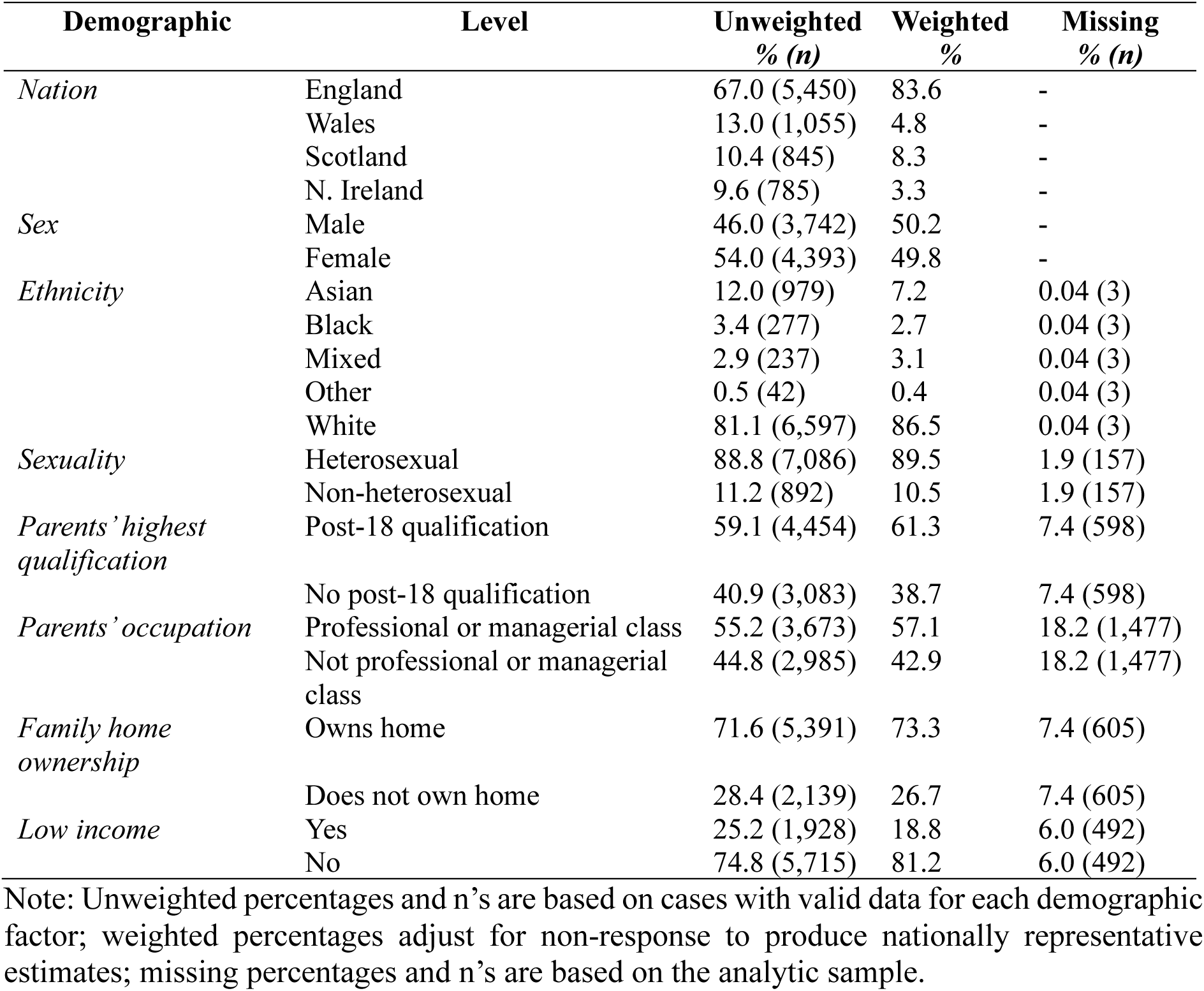
Analytic sample demographics (n = 8,135).

### Adjusted relative risks

*Table 2* presents the prevalences and adjusted relative risks (aRRs) for each health exposure after appropriate weighting and adjustment for demographic factors. The aRRs are also shown in *Figure 2*. *Supplementary Material C* provides the prevalence of each demographic factor and health exposure for the age 23 NEET and non-NEET status groups separately. *Supplementary Material D* presents the aRRs without using imputed data. *Supplementary Material E* presents the equivalent RRs but without adjustment for demographic factors.

**Figure 2.**
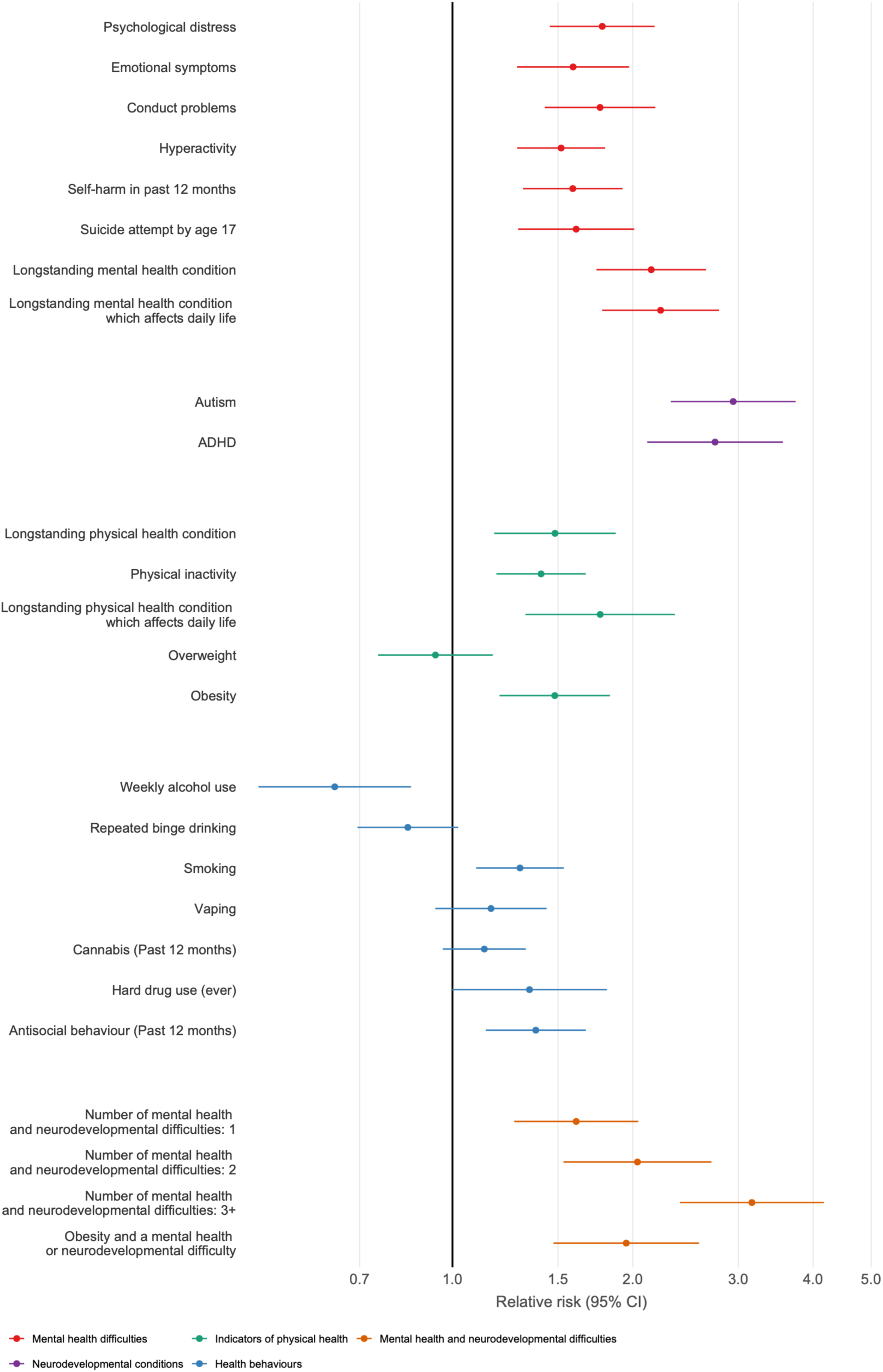
Relative risks of being NEET at age 23 adjusting for demographics (nation, sex, ethnicity, sexuality, socioeconomic background). 95% CI = 95% confidence intervals. N = 8,135, imputed data.

**Table 2.** Prevalences of health exposures in the analytic sample accompanied by corresponding relative risks and population attributable fractions of being NEET at age 23 (n = 8,135) using imputed data.

| <u>Exposure</u> | <u>Unadjusted prevalences</u> |  |  | <u>Adjusted, weighted, imputed</u> |  | <u>Population attributable fractions</u><br>*PAF*<br>(%)<br>*p<sub>c</sub>* |  |
| --- | --- | --- | --- | --- | --- | --- | --- |
|  | Unweighted | Weighted | Missing | Relative risks |  |  |  |
|  |  |  |  | *aRR* | *95% CI* |  |  |
|  | % (*n*) | % | % (*n*) |  |  |  |  |
| <u>Mental health difficulties</u> |  |  |  |  |  |  |  |
| *Psychological distress* | 16.7 (1,334) | 15.9 | 1.6 (133) | 1.78* | 1.45, 2.18 | 11.90 | 27.19 |
| *Conduct problems* | 4.9 (393) | 5.0 | 2.0 (162) | 1.76* | 1.43, 2.18 | 4.68 | 10.8 |
| *Emotional symptoms* | 14.7 (1,173) | 14.1 | 2.0 (162) | 1.59* | 1.28, 1.97 | 7.88 | 21.24 |
| *Hyperactivity* | 14.3 (1,138) | 14.3 | 2.0 (162) | 1.52* | 1.28, 1.80 | 7.67 | 22.47 |
| *Self-harm in past 12 months* | 24.2 (1,926) | 23.9 | 2.0 (173) | 1.59* | 1.31, 1.92 | 13.06 | 35.27 |
| *Suicide attempt by age 17* | 7.5 (593) | 7.4 | 2.1 (180) | 1.61* | 1.29, 2.01 | 5.53 | 14.61 |
| *Longstanding mental health condition...* | 6.2 (499) | 6.9 | 1.1 (93) | 2.15* | 1.74, 2.65 | 8.97 | 16.79 |
| *... which affects daily life* | 5.3 (427) | 6.0 | 1.1 (93) | 2.23* | 1.78, 2.79 | 8.44 | 15.32 |
| <u>Childhood neurodevelopmental conditions</u> |  |  |  |  |  |  |  |
| *Autism* | 3.6 (294) | 4.2 | 0.2 (14) | 2.94* | 2.31, 3.74 | 9.76 | 14.79 |
| *ADHD* | 2.6 (210) | 2.9 | 0.2 (14) | 2.74* | 2.11, 3.56 | 6.54 | 10.29 |
| <u>Indicators of physical health</u> |  |  |  |  |  |  |  |
| *Overweight* | 19.5 (1,489) | 19.2 | 6.0 (485) | 0.94 | 0.75, 1.17 | n/a | 17.81 |
| *Obesity* | 10.8 (829) | 10.2 | 6.0 (485) | 1.48** | 1.20, 1.83 | 5.96 | 18.34 |
| *Longstanding physical health condition...* | 7.9 (638) | 7.9 | 1.1 (93) | 1.48** | 1.17, 1.87 | 4.09 | 12.55 |
| *... which affects daily life* | 5.0 (406) | 5.1 | 1.1 (93) | 1.76** | 1.32, 2.35 | 4.33 | 10.0 |
| *No physical activity* | 24.7 (1,984) | 22.9 | 1.1 (91) | 1.41** | 1.18, 1.67 | 9.44 | 32.69 |
| <u>Health behaviours</u> |  |  |  |  |  |  |  |
| *Weekly alcohol use* | 11.6 (929) | 13.3 | 1.9 (154) | 0.64** | 0.47, 0.85 | n/a | 7.41 |
| *Repeated binge drinking* | 30.3 (2,415) | 32.3 | 1.9 (157) | 0.84 | 0.69, 1.02 | n/a | 25.53 |
| *Smoking* | 18.4 (1,467) | 20.1 | 1.9 (160) | 1.30** | 1.10, 1.53 | 5.98 | 26.18 |
| *Vaping* | 10.6 (842) | 11.0 | 1.9 (161) | 1.16 | 0.94, 1.44 | n/a | 13.85 |
| *Cannabis use in the past 12 months* | 27.5 (2,192) | 30.1 | 1.8 (152) | 1.13 | 0.96, 1.33 | n/a | 32.79 |
| *Hard drug use ever* | 8.2 (653) | 9.6 | 1.7 (141) | 1.34 | 1.00, 1.81 | n/a | 12.1 |

| <u>Exposure</u> | <u>Unadjusted prevalences</u> |  |  | <u>Adjusted, weighted, imputed</u> |  | Population attributable fractions<br>*PAF*<br>(%)<br>*p<sub>c</sub>* |  |
| --- | --- | --- | --- | --- | --- | --- | --- |
|  | Unweighted | Weighted | Missing | Relative risks |  |  |  |
|  | % (*n*) | % | % (*n*) | *aRR* | 95% *CI* |  |  |
| *Antisocial behaviour in the past 12 months* | 10.5 (842) | 11.4 | 1.6 (133) | 1.38** | 1.14, 1.67 | 4.28 | 15.61 |
| <u>Mental health and neurodevelopmental difficulties</u> |  |  |  |  |  |  |  |
| *1 difficulty* | 19.9 (1,622) | 19.9 | 0.01 (1) | 1.61* | 1.27, 2.04 | 8.82 | 23.32 |
| *2 difficulties* | 9.5 (769) | 9.2 | 0.01 (1) | 2.04* | 1.53, 2.71 | 7.39 | 14.52 |
| *3+ difficulties* | 6.0 (491) | 6.0 | 0.01 (1) | 3.16* | 2.40, 4.17 | 11.21 | 16.39 |
| *Obesity and a difficulty* | 4.5 (348) | 4.4 | 6.0 (486) | 1.95* | 1.47, 2.58 | 5.32 | 10.91 |
| NEET at age 17 | 3.2 (256) | 3.1 | 1.1 (91) | 2.96* | 2.4, 3.64 | 8.13 | 12.29 |

Note: Unweighted n's and prevalences are based on cases with valid data for each exposure.
Weighted prevalence, relative risks and PAFs adjust for non-response to produce nationally representative estimates. Each relative risk was calculated independently. Adjusted relative risks controlled for nation, sex, ethnicity, sexuality and socioeconomic background.
Reference categories are not reporting the exposure, except for overweight and obesity, where the reference is being not overweight. All p-values are corrected using the Holm procedure.
PAF estimates are not provided where the corresponding aRR was below 1 or if the association was non-significant after the Holm procedure. PAFs were calculated based on non-imputed $p_c$ .
Abbreviations: 95% CI = 95% confidence intervals; ADHD = attention deficit hyperactivity disorder; aRR = adjusted relative risks; PAF = population attributable fractions; $p_c$ = weighted prevalence of exposure among NEET cases.
Significance levels: \* = $p < .001$ ; \*\* = $p < .05$ .

Regarding adolescent mental health difficulties, a self-reported longstanding mental health condition more than doubled the risk of being NEET (aRR = 2.15, 95% CI = 1.74, 2.65), with a slightly stronger association among those reporting that it affected their daily life (aRR = 2.23, 95% CI = 1.78, 2.79). Psychological distress (aRR = 1.78, 95% CI = 1.45, 2.18), conduct problems (aRR = 1.76, 95% CI = 1.43, 2.18), emotional symptoms (aRR = 1.59, 95% CI = 1.28, 1.97), hyperactivity (aRR = 1.52, 95% CI = 1.28, 1.80), self-harm in the past 12 months (aRR = 1.59, 95% CI = 1.31, 1.92), and a suicide attempt by age 17 (aRR = 1.61, 95% CI = 1.29, 2.01) were associated with a higher risk of being NEET at age 23 by at least 50%.

For childhood neurodevelopmental conditions, cohort members with an autism diagnosis (aRR = 2.94, 95% CI = 2.31, 3.74) or ADHD diagnosis (aRR = 2.74, 95% CI = 2.11, 3.56) were almost three times more likely to be NEET by age 23 than their peers.

Regarding indicators of physical health in adolescence, obesity was associated with an approximately 50% higher risk of being NEET by age 23 (aRR = 1.48, 95% CI = 1.20, 1.83). A longstanding physical health condition alone was associated with later NEET status (aRR = 1.48, 95% CI = 1.17, 1.87) and a condition that affected daily life was associated with a higher risk by approximately 75% (aRR = 1.76, 95% CI = 1.32, 2.35). No physical activity was associated with a higher risk of later NEET status (aRR = 1.41, 95% CI = 1.18, 1.67).

Regarding adolescent health behaviours, unexpectedly, weekly alcohol use was associated with a lower risk of being NEET at age 23 (aRR = 0.64, 95% CI = 0.47, 0.85). The adolescent health behaviours most strongly associated with an increased likelihood of later NEET status were smoking (aRR = 1.30, 95% CI = 1.10, 1.53) and self-reported antisocial behaviour in the past 12 months (aRR = 1.38, 95% CI = 1.14, 1.67).

Regarding mental health and neurodevelopmental difficulties, having any difficulty was associated with a higher risk of being NEET at age 23 compared to having none (aRR = 1.61, 95% CI = 1.27, 2.04). Having two difficulties more than doubled the risk (aRR = 2.04, 95% CI = 1.53, 2.71), while having at least three tripled it (aRR = 3.16, 95% CI = 2.40, 4.17). Adolescent obesity combined with a mental health or neurodevelopmental difficulty was associated with approximately double the risk of being NEET at age 23 (aRR = 1.95, 95% CI = 1.47, 2.58).

*Supplementary Material F* describes two sets of sensitivity analyses. First, cohort members who were NEET at age 17 were almost three times as likely to also be NEET at age 23 (aRR = 2.96, 95% CI = 2.40, 3.64). To reduce the likelihood that effects were being driven by the unique characteristics of cohort members who were already NEET by age 17, as a sensitivity analysis, adjusted relative risks were also calculated separately for the cohort members who were not NEET by age 17. The overall pattern was similar. Second, analyses distinguishing unemployment from economic inactivity in the non-imputed data suggested that health effects were largely concentrated among economically inactive young adults, with weaker and less consistent associations among those actively seeking work.

### Population attributable fractions (PAFs)

The four adolescent health exposures with the largest estimated PAFs for young adult NEET prevalence were self-harm in the past 12 months (13.06%), psychological distress (11.90%), autism (9.76%) and no physical activity (9.44%). PAF estimates are reported on the right side of *Table 2*.

### Analyses by sex

*Table 3* provides the aRRs and PAFs for being NEET at age 23 stratified by sex. *Supplementary Material G* presents the same analyses without using imputed data. aRRs were generally larger for women than men. Interaction terms in the non-imputed data suggested that the association between hyperactivity at age 17 and NEET status at age 23 differed by sex (*p* = .004), but this association did not persist in the imputed data. PAFs were generally larger for women than men for almost all mental health difficulties, except for autism (4.14% and 14.89%) and ADHD (2.95% and 9.77%). The largest PAFs were psychological distress (18.19%) and self-harm in the past 12 months (17.60%) for women, and autism (14.89%) and no physical activity (11.56%) for men.

**Table 3.** Weighted health exposure adjusted relative risks and population attributable fractions of being NEET at age 23 (n = 8,135), divided by sex, based on the imputed dataset.

| <u>Exposure</u> | <u>Adjusted relative risks</u> |  |  |  | <u>Population attributable fractions</u> |  |  |  |
| --- | --- | --- | --- | --- | --- | --- | --- | --- |
|  | Female |  | Male |  | Female |  | Male |  |
|  | *aRR* | 95%<br>*CI* | *aRR* | 95%<br>*CI* | *PAF*<br>(%) | *p<sub>c</sub>* | *PAF*<br>(%) | *p<sub>c</sub>* |
| <u>Mental health difficulties</u> |  |  |  |  |  |  |  |  |
| *Psychological distress* | 1.88* | 1.47, 2.40 | 1.64** | 1.21, 2.21 | 18.19 | 38.97 | 6.58 | 16.93 |
| *Conduct problems* | 1.98* | 1.45, 2.72 | 1.65** | 1.27, 2.15 | 4.87 | 9.82 | 4.62 | 11.67 |
| *Emotional symptoms* | 1.61** | 1.26, 2.06 | 1.55 | 1.11, 2.15 | 12.57 | 33.25 | n/a | 10.57 |
| *Hyperactivity* | 1.67* | 1.33, 2.10 | 1.39 | 1.07, 1.79 | 9.71 | 24.14 | n/a | 20.99 |
| *Self-harm in past 12 months* | 1.66** | 1.26, 2.20 | 1.52** | 1.19, 1.95 | 17.60 | 44.22 | 9.45 | 27.51 |
| *Suicide attempt by age 17* | 1.57** | 1.19, 2.06 | 1.64 | 1.16, 2.31 | 7.45 | 20.6 | n/a | 9.48 |
| *Longstanding mental health condition...* | 2.09* | 1.51, 2.89 | 2.24* | 1.66, 3.01 | 11.03 | 21.2 | 7.21 | 13.03 |
| *... which affects daily life* | 2.20* | 1.58, 3.05 | 2.30* | 1.69, 3.14 | 10.27 | 18.85 | 6.97 | 12.31 |
| <u>Childhood neurodevelopmental conditions</u> |  |  |  |  |  |  |  |  |
| *Autism* | 2.53* | 1.68, 3.81 | 3.25* | 2.48, 4.25 | 4.14 | 6.85 | 14.89 | 21.51 |
| *ADHD* | 2.81* | 1.79, 4.41 | 2.83* | 2.16, 3.70 | 2.95 | 4.58 | 9.77 | 15.12 |
| <u>Indicators of physical health</u> |  |  |  |  |  |  |  |  |
| *Overweight* | 0.90 | 0.68, 1.20 | 0.97 | 0.73, 1.31 | n/a | 18.07 | n/a | 17.61 |
| *Obesity* | 1.42 | 1.03, 1.96 | 1.54** | 1.16, 2.05 | n/a | 18.69 | 6.33 | 18.05 |
| *Longstanding physical health condition...* | 1.38 | 0.96, 1.99 | 1.60 | 1.14, 2.23 | n/a | 13.87 | n/a | 11.42 |
| *... which affects daily life* | 1.62 | 1.09, 2.41 | 1.96** | 1.30, 2.94 | n/a | 11.64 | 4.21 | 8.60 |
| *No physical activity* | 1.21 | 0.95, 1.54 | 1.65* | 1.29, 2.10 | n/a | 36.61 | 11.56 | 29.34 |
| <u>Health behaviours</u> |  |  |  |  |  |  |  |  |
| *Weekly alcohol use* | 0.78 | 0.53, 1.15 | 0.56 | 0.36, 0.86 | n/a | 6.88 | n/a | 7.87 |
| *Repeated binge drinking* | 0.94 | 0.73, 1.20 | 0.76 | 0.57, 1.01 | n/a | 25.95 | n/a | 25.17 |
| *Smoking* | 1.47** | 1.14, 1.89 | 1.15 | 0.88, 1.48 | 9.72 | 30.4 | n/a | 22.5 |
| *Vaping* | 1.23 | 0.85,<br>1.77 | 1.11 | 0.85,<br>1.46 | n/a | 12.1 | n/a | 15.38 |
| *Cannabis use in the past*<br>*12 months* | 1.14 | 0.90,<br>1.44 | 1.12 | 0.89,<br>1.40 | n/a | 31.99 | n/a | 33.48 |
| *Hard drug use ever* | 1.36 | 0.94,<br>1.97 | 1.33 | 0.93,<br>1.91 | n/a | 10.32 | n/a | 13.65 |
| *Antisocial behaviour in the*<br>*past 12 months* | 1.44 | 1.01,<br>2.06 | 1.32 | 1.01,<br>1.73 | n/a | 13.13 | n/a | 17.77 |
| <u>Mental health and</u><br><u>neurodevelopmental</u><br><u>difficulties</u> |  |  |  |  |  |  |  |  |
| *1 difficulty* | 1.36 | 0.94,<br>1.95 | 1.88* | 1.44,<br>2.46 | n/a | 22.8 | 11.11 | 23.76 |
| *2 difficulties* | 1.99** | 1.38,<br>2.86 | 2.06** | 1.40,<br>3.03 | 8.18 | 16.44 | 6.62 | 12.87 |
| *3+ difficulties* | 2.94* | 2.10,<br>4.10 | 3.27* | 2.27,<br>4.71 | 13.07 | 19.81 | 9.35 | 13.47 |
| *Obesity and a difficulty* | 1.81** | 1.22,<br>2.68 | 2.10* | 1.47,<br>3.00 | 5.51 | 12.33 | 5.11 | 9.77 |
| <u>NEET at age 17</u> | 2.85* | 2.12,<br>3.84 | 3.08* | 2.27,<br>4.17 | 8.20 | 12.62 | 8.10 | 12 |

Note: Weighted prevalence rates, relative risks and PAFs adjust for non-response to produce nationally representative estimates. Each relative risk was calculated independently. Adjusted relative risks controlled for nation, sex, ethnicity, sexuality and socioeconomic background. Reference categories are not reporting the condition or behaviour, except for overweight and obesity, where the reference is being not overweight, including being underweight. All p-values are corrected using the Holm procedure. PAF estimates are not provided where the corresponding aRR was below 1 or if the association was non-significant after the Holm procedure.
Abbreviations: 95% CI = 95% confidence intervals; ADHD = attention deficit hyperactivity disorder; aRR = adjusted relative risks; PAF = population attributable fractions; $p_c$ = weighted prevalence of exposure among NEET cases. Significance levels: \* = $p < .001$ ; \*\* = $p < .05$ .

## Discussion

These analyses drew on a large, broadly nationally representative UK cohort of young adults to examine whether a wide range of theoretically informed health exposures during adolescence were associated with NEET status in early adulthood six years later for Gen Z. The findings suggest that adolescent mental health difficulties and childhood neurodevelopmental conditions are particularly associated with NEET status at age 23, even after adjusting for key demographic factors. However, these associations are not to be interpreted as direct causal pathways and are likely to be confounded by other factors that may affect both health and NEET status. Instead, these associations help identify young people who might benefit from additional support as they transition into education or the workforce. Our study also demonstrates notable variation between health exposures in their strength of association and prevalence, with the PAF estimates highlighting the need to understand risk associated with varied factors and consider these in the context of population prevalence.

Adolescent mental health difficulties and childhood neurodevelopmental conditions were consistently associated with NEET status at age 23. All adolescent mental health difficulties were associated with a higher risk of later NEET status. Reporting a longstanding mental health condition, especially one that affected cohort members’ daily life for more than six months, doubled NEET risk. Effects were stronger again for reports of childhood neurodevelopmental conditions: autism or ADHD diagnoses almost tripled the risk of being NEET in young adulthood. Given the increased prevalence of these diagnoses in Gen Z young people, this should be of particular concern for policymakers (29).

Young adults who are NEET are frequently characterised by the accumulation of multiple mental health and neurodevelopmental difficulties. We therefore aimed to account for the cumulative nature of risk and how co-occurring earlier difficulties were associated with later adult NEET status. Compared to cohort members who reported no mental health difficulties or childhood neurodevelopmental conditions, having a single difficulty increased the risk of being NEET at age 23 by approximately 60%. Approximately 6% of the sample recorded three or more difficulties at age 17, which tripled their risk of being NEET at age 23.

Indicators of physical health in adolescence were also linked to later NEET status. Longstanding conditions affecting daily life were associated with a 76% higher risk. Obesity, but not being overweight, was associated with a 48% higher risk of being NEET at age 23. An adolescent’s weight could serve as a risk factor for exclusion from the labour market as a young adult through a number of potential pathways, including weight-based stigma, discrimination and obesity-related health problems. Physical inactivity was examined as a separate indicator of physical health and was also associated with being NEET at age 23. Cohort members with co-occurring obesity and mental health or neurodevelopmental difficulties had notably elevated risks of being NEET which suggests that accumulating multiple health challenges may be more relevant to later NEET status than individual exposures alone.

PAFs highlighted that common adolescent health problems may have a greater population impact on later NEET status than rarer but more severe conditions. For example, self-harming in the past 12 months and physical inactivity had some of the largest PAFs, despite not having the largest aRRs. Although mental health difficulties and childhood neurodevelopmental conditions had notably larger aRRs on average than indicators of physical health, there was a greater balance between the four sets of exposures in the list of exposures with the largest PAFs. As this is an observational study, we cannot establish causality. The PAFs should therefore be treated as descriptive estimates that combine the prevalence of each health exposure with its adjusted association, rather than as estimates of the proportion of NEET cases that would be prevented if the exposure were eliminated. These findings may help prioritise targets for interventions that balance severity and prevalence for Gen Z young people.

Analyses were also conducted stratified by sex. Although baseline NEET rates did not themselves show a significant sex difference, several associations were comparatively larger among women than men, although interaction tests did not provide clear evidence of sex differences.

These analyses have a number of limitations. First, young people experiencing difficult circumstances or economic challenges are less likely to be successfully contacted by longitudinal studies; therefore, the prevalence of NEET in young people may be underestimated. Although the survey weights are intended to account for differential attrition, selection bias is still possible if the weights were mis-specified by not including unmeasured factors related to NEET status specifically. Second, this is an observational study and there are likely unmeasured confounding variables that relate to health exposure and NEET status (for example, parental mental health, prior academic achievement and engagement, genetics) that are not included in models; hence, these estimates should not be interpreted as causal. Third, we chose to dichotomise some ordinal variables to ease interpretability and to match cut-offs used in the literature. A limitation of this decision is that it reduces resolution and ignores variation between collapsed categories, which may have affected results. Fourth, analyses of potential selection bias suggested that the analytic sample was disproportionately White, from a higher socioeconomic background and not already NEET at age 17 compared to participants excluded from the analytic sample In addition, sensitivity analyses suggested that the unweighted prevalence of NEET at age 23 was 13.5% in the overall MCS sample at age 23, compared to 12.3% in the analytic sample, suggesting slight additional selection bias. Fifth, our exposure variables were self-reported and therefore subject to measurement error from recall bias or social desirability bias.

As the MCS cohort ages further, future research should examine the longer-term consequences of health and being NEET, and how they interact. Consistent with the idea of a scarring effect, being NEET at age 17 tripled the risk of being NEET at age 23. As these analyses mainly use exposure data from age 17, it is possible education disengagement that had already occurred or early experiences of the transition to the labour market had affected cohort members’ health before age 17 (i.e. reverse causality). A limitation of the data is that economic activity was only measured once at a single timepoint, making it difficult to differentiate between short periods of NEET status and prolonged disengagement from the labour market. Further research that more frequently measures both economic activity and health experiences could help distinguish temporary disengagement during educational transitions from chronic NEET status and better identify the pathways linking health and labour market outcomes. Further comorbidities should also be considered: we intended to examine co-occurrence between obesity and self-reported longstanding health conditions, but analyses would have been underpowered. In addition, the role of inequalities as a moderator was not the focus of these analyses, but future analyses of the MCS could consider how NEET status can be transmitted between generations and what mechanisms explain individual differences in the strengths of those links. Similarly, associations are likely to vary by socioeconomic background, ethnicity, sexuality, disability and region, which future research should investigate.

In conclusion, these analyses show that for Gen Z, specific health exposures in adolescence may be among several factors linked to a higher likelihood of NEET status in young adulthood. The findings help identify young people who may benefit from targeted health and employment support, while highlighting the importance of early intervention to prevent health problems translating into labour market exclusion. More broadly, they suggest that being NEET is not only an economic issue but also a health issue, requiring coordinated action across education, health and employment services. These findings suggest that the risk of being NEET falls disproportionately on young people who are already disadvantaged by their health. It is likely that continued economic and societal shocks will disproportionately affect the socioeconomically disadvantaged members of Gen Z as they have the least resources to adjust to changes. Therefore, it is essential that policy conversations regarding the relationships between health and economic activity also address the structural inequalities that are still prevalent in society.

## Data Availability

The data that support the findings of this study are available from the UK Data Service. Restrictions apply to the availability of these data, which were used under license for this study.

https://datacatalogue.ukdataservice.ac.uk

## Declarations

### Ethics approval and consent to participate

Ethical approval for the Millennium Cohort Study was provided by the NHS Multi-Centre Ethics Committee and for these analyses by the UCL Institute of Education research ethics committee (REC 2428). Cohort members provided active consent to be included in the study.

### Consent for publication

Not applicable.

### Authors’ contributions

DK: Conceptualisation, Methodology, Formal Analysis, Data Curation, Writing-Original Draft, Visualisation; JW-Writing-Review & Editing; PP: Conceptualisation, Methodology, Writing-Review & Editing, Supervision, Project Administration. All authors read and approved the final manuscript.

### Artificial intelligence use

DK used Microsoft 365 Copilot in October 2026 to assist with language editing, proof-reading and code checking. All text and code were reviewed and verified by the authors, who take full responsibility for the final content.

## Acknowledgements

Thank you to the participants of the Millennium Cohort Study and their family members for their time. These findings would not have been possible without their invaluable contributions. We are very grateful for their ongoing commitment to the study.

## List of Abbreviations

ADHD: attention deficit hyperactivity disorder
aRR: adjusted relative risk
BMI: body mass index
CI: confidence interval
Gen Z: Generation Z
MCS: Millennium Cohort Study
NEET: not in education, employment or training
NHS: National Health Service
OECD: Organisation for Economic Co-operation and Development
PAF: population attributable fraction
p_c_: weighted prevalence of exposure among NEET cases
RR: relative risk
SDQ: Strengths and Difficulties Questionnaire
UK: United Kingdom

## References

Office for National Statistics. Young people not in Education, Employment or Training (NEET), UK: February 2025. Newport: Office for National Statistics; 2025.

Chandler RF, Lozada ARS. Health status among NEET adolescents and young adults in the United States, 2016–2018. SSM - Population Health. 2021;14:100814.

Gunnes M, Thaulow K, Kaspersen SL, Jensen C, Ose SO. Young adults not in education, employment, or training (NEET): a global scoping review. BMC Public Health. 2025;25(1):3394.

Rodwell L, Romaniuk H, Nilsen W, Carlin JB, Lee KJ, Patton GC. Adolescent mental health and behavioural predictors of being NEET: a prospective study of young adults not in employment, education, or training. Psychological Medicine. 2018;48(5):861–71.

Gariepy G, Danna SM, Hawke L, Henderson J, Iyer SN. The mental health of young people who are not in education, employment, or training: a systematic review and meta-analysis. Social Psychiatry and Psychiatric Epidemiology. 2022;57(6):1107–21.

Hammerton G, Murray J, Maughan B, Barros FC, Goncalves H, Menezes AMB, et al. Childhood behavioural problems and adverse outcomes in early adulthood: A comparison of Brazilian and British birth cohorts. Journal of Developmental and Life-Course Criminology. 2019;5(4):517–35.

Mars B, Heron J, Crane C, Hawton K, Lewis G, Macleod J, et al. Clinical and social outcomes of adolescent self harm: population based birth cohort study. BMJ. 2014;349:g5954.

Lindblad V, Ravn RL, Gaardsted PS, Hansen LEM, Lauritzen FF, Melgaard D. Beyond the mind: Understanding the influence of mental health on youth NEET status in Europe-A systematic literature review. J Adolescence. 2024;96(7):1428–44.

Agnew-Blais JC, Polanczyk GV, Danese A, Wertz J, Moffitt TE, Arseneault L. Young adult mental health and functional outcomes among individuals with remitted, persistent and late-onset ADHD. The British Journal of Psychiatry. 2018;213(3):526–34.

Lindblad V, Kragholm KH, Gaardsted PS, Hansen LEM, Lauritzen FF, Melgaard D. From illness to inactivity: Exploring the influence of physical diseases on youth Not in Education, Employment, or Training status in Europe: A systematic literature review. J Adolescence. 2024;96(8):1695–712.

Kansra AR, Lakkunarajah S, Jay MS. Childhood and adolescent obesity: A review. Frontiers in Pediatrics. 2020;8:581461.

Baggio S, Iglesias K, Deline S, Studer J, Henchoz Y, Mohler-Kuo M, et al. Not in education, employment, or training status among young Swiss men. Longitudinal associations with mental health and substance use. Journal of Adolescent Health. 2015;56(2):238–43.

Villadsen A, Fitzsimons E. Substance use and addictive behaviours. London: Centre for Longitudinal Studies; 2026.

Rahmani H, Groot W, Rahmani AM. Unravelling the NEET phenomenon: a systematic literature review and meta-analysis of risk factors for youth not in education, employment, or training. International Journal of Adolescence and Youth. 2024;29(1):2331576.

University of London, UCL Social Research Institute, Centre for Longitudinal Studies. Millennium Cohort Study: Sweep 8, Age 23, 2023-2025. 2 ed: UK Data Service; 2026.

University of London IoE, Centre for Longitudinal Studies. Millennium Cohort Study: Age 17, Sweep 7, 2018. 2 ed: UK Data Service; 2024.

Beltz AM, Kelly DP, Berenbaum SA. Sex differences in brain and behavioral development. In: Rubenstein J, Rakic P, editors. Neural Circuit and Cognitive Development. 2nd ed2020. p. 585–638.

Ipsos. Millennium Cohort Study Eighth Sweep (MCS8): Technical Report. London: Centre for Longitudinal Studies, UCL Social Research Institute; 2025.

Ipsos MORI. Millennium Cohort Study Seventh Sweep (MCS7): Technical Report. London: Centre for Longitudinal Studies, UCL Institute of Education; 2019.

Office for National Statistics. A guide to labour market statistics 2020 [updated 16 June 2020. Available from: https://www.ons.gov.uk/employmentandlabourmarket/peopleinwork/employmentandemployeetypes/methodologies/aguidetolabourmarketstatistics.

Kessler RC, Barker PR, Colpe LJ, Epstein JF, Gfroerer JC, Hiripi E, et al. Screening for serious mental illness in the general population. Archives of General Psychiatry. 2003;60(2):184–9.

Goodman R, Meltzer H, Bailey V. The Strengths and Difficulties Questionnaire: a pilot study on the validity of the self-report version. European Child & Adolescent Psychiatry. 1998;7(3):125–30.

University of London, Institute of Education, Centre for Longitudinal Studies. Millennium Cohort Study: Age 5, Sweep 3, 2006. 9 ed: UK Data Service; 2024.

University of London, Institute of Education, Centre for Longitudinal Studies. Millennium Cohort Study: Age 7, Sweep 4, 2008. 10 ed: UK Data Service; 2025.

University of London, Institute of Education, Centre for Longitudinal Studies. Millennium Cohort Study: Age 11, Sweep 5, 2012. 6 ed: UK Data Service; 2024.

University of London, Institute of Education, Centre for Longitudinal Studies. Millennium Cohort Study: Age 14, Sweep 6, 2015. 8 ed: UK Data Service; 2025.

Mansournia MA, Altman DG. Population attributable fraction. BMJ. 2018;360:k757.

Van Buuren S, Groothuis-Oudshoorn K. mice: Multivariate imputation by chained equations in R. Journal of Statistical Software. 2011;45:1–67.

Fonagy P, Baird G, Wessely S. Independent review into mental health conditions, ADHD and autism: Interim report. Department of Health and Social Care; 2026.

Kessler RC, Andrews G, Colpe LJ, Hiripi E, Mroczek DK, Normand SL, et al. Short screening scales to monitor population prevalences and trends in non-specific psychological distress. Psychological Medicine. 2002;32(6):959–76.

Goodman R. Psychometric properties of the strengths and difficulties questionnaire. Journal of the American Academy of Child & Adolescent Psychiatry. 2001;40(11):1337–45.

Bélair M-A, Kohen DE, Kingsbury M, Colman I. Relationship between leisure time physical activity, sedentary behaviour and symptoms of depression and anxiety: evidence from a population-based sample of Canadian adolescents. BMJ Open. 2018;8(10):e021119.

Silins E, Horwood LJ, Najman JM, Patton GC, Toumbourou JW, Olsson CA, et al. Adverse adult consequences of different alcohol use patterns in adolescence: an integrative analysis of data to age 30 years from four Australasian cohorts. Addiction. 2018;113(10):1811–25.

Viner RM, Taylor B. Adult outcomes of binge drinking in adolescence: findings from a UK national birth cohort. Journal of Epidemiology and Community Health. 2007;61(10):902–7.

