## Supplementary Files for "Adolescent health and not in education, employment or training (NEET) in young adulthood: Evidence from a UK prospective longitudinal study"

**Supplementary Material A: Characteristics of cohort members who participated in the age 17 sweep who were retained and not retained in the analytic sample**

| Demographic | Level | Overall (n = 10,726) n (%) | Retained (n = 8,135) n (%) | Not retained (n = 2,591) n (%) |
|---|---|---|---|---|
| *Nation* | England | 7,193 (84) | 5,450 (84) | 1,743 (84) |
| | Wales | 1,421 (5.3) | 1,055 (4.8) | 366 (6.5) |
| | Scotland | 1,119 (7.3) | 845 (7.6) | 274 (6.6) |
| | N. Ireland | 993 (3.2) | 785 (3.3) | 208 (2.9) |
| *Sex* | Male | 5,359 (51) | 3,742 (46) | 1,617 (62) |
| | Female | 5,367 (49) | 4,393 (54) | 974 (38) |
| *Ethnicity* | Asian | 1,280 (6.4) | 979 (6.6) | 301 (5.9) |
| | Black | 384 (3.2) | 277 (2.9) | 107 (4) |
| | Mixed | 322 (3.6) | 237 (3.3) | 85 (4.1) |
| | Other | 58 (0.6) | 42 (0.3) | 16 (1.1) |
| | White | 8,676 (86) | 6,597 (87) | 2,079 (85) |
| | *Missing* | 6 (0.1) | 3 (0.04) | 3 (0.1) |
| *Sexuality* | Heterosexual | 8,806 (90) | 7,086 (89) | 1,720 (91.2) |
| | Non-heterosexual | 1,038 (10) | 892 (11) | 146 (8.8) |
| | *Missing* | 882 (8.2) | 157 (1.9) | 725 (28) |
| *Parents' highest qualification* | Post-18 qualification | 5,483 (56) | 4,454 (59) | 1,029 (46) |
| | No post-18 qualification | 4,193 (44) | 3,083 (41) | 1,110 (54) |
| | *Missing* | 1,050 (9.8) | 598 (7.4) | 452 (17.4) |
| *Parents' occupation* | Professional or managerial class | 4,512 (56) | 3,673 (58) | 839 (48) |
| | Not professional or managerial class | 3,880 (44) | 2,985 (42) | 895 (52) |
| | *Missing* | 2,334 (21.8) | 1,477 (18.2) | 857 (33.1) |
| *Family home ownership* | Owns home | 6,712 (68) | 5,391 (72) | 1,321 (57) |
| | Does not own home | 2,963 (32) | 2,139 (28) | 824 (43) |
| | *Missing* | 1,051 (9.8) | 605 (7.4) | 446 (17.2) |
| *Low income* | Yes | 2,705 (23) | 1,928 (19) | 777 (34) |
| | No | 7,130 (77) | 5,715 (81) | 1,415 (66) |
| | *Missing* | 891 (8.3) | 492 (6) | 399 (15.4) |
| *NEET at age 17* | Yes | 361 (3.9) | 256 (3) | 105 (6.9) |
| | No | 9,609 (96) | 7,788 (97) | 1,821 (93) |
| | *Missing* | 756 (7) | 91 (1.1) | 665 (25.7) |

Note: ns are unweighted, percentages are weighted, with the exception of percentages for the missing data row, which are unweighted. Weighted percentages are based on cases with valid data for each demographic factor and adjust for non-response to produce nationally representative estimates. 29 participants were not included in analyses of data of the overall sample at the age 17 sweep due to their data being unavailable or excluded for disclosure reasons.

**Supplementary Material B: Further details on the outcome and exposures**

*NEET status at age 23.* Cohort members in paid work, self-employment, in education, on apprenticeships, government training schemes, maternity leave or waiting to start a job were classified as not NEET (0). Those unemployed, sick or disabled, caring for home or family, on a career break, seeking work or in unpaid or volunteer work were classified as NEET (1). Unclassifiable responses were coded as missing.

*Mental health difficulties.* Cohort members reported a range of mental health symptoms and conditions at age 17.

1. *Psychological distress.* The six-item Kessler Scale (30) measured non-specific psychological distress over the past 30 days. Items were scored from 0-4 and summed (range 0-24). Cohort members scoring ≥13 were classified as experiencing high psychological distress (21). Missing data were handled using mean imputation where at least three items were completed; responses were removed if fewer than three items were completed.
2. *Conduct problems, emotional symptoms and hyperactivity.* Three subscales from the self-report version of the SDQ (22), which has strong psychometric properties (31), were used to measure conduct problems, emotional symptoms and hyperactivity. Items were scored from 0-2 and summed (range 0-10 for each subscale). Following standard scoring procedures (www.sdqinfo.org), participants scoring in the high range (≥7 for conduct problems and hyperactivity; ≥5 for emotional symptoms) were classified as the corresponding mental health difficulty. Mean imputation was applied where sufficient responses were available; responses were otherwise removed.
3. *12-month self-harm; suicide attempt by age 17.* Cohort members reporting self-harm in the past year (via six items regarding different means) were classified

as having self-harmed. Suicide attempt by age 17 was assessed with a single follow-up item.

4. *Longstanding mental health condition.* Cohort members who reported a physical or mental health condition or illness expected to last 12 months or more were classified as having a longstanding mental health condition if at least one of their conditions concerned their mental health. A separate series of two items identified conditions reported to impact daily activities for more than six months. Cohort members could report multiple longstanding conditions, including conditions which did not concern mental health, so the question regarding the impact of their conditions on their daily activities may not have reflected mental health conditions alone.

*Childhood neurodevelopmental conditions.* Parent-reported diagnoses of autism and ADHD were collected at ages 5, 7, 11 and 14. At ages 11 and 14, only parents who had not previously reported a diagnosis were asked about new diagnoses. Because reports were sometimes inconsistent across waves, cohort members were classified as having a condition if it was reported at any wave.

*Indicators of physical health.* Cohort members responded to items at age 17 regarding:

1. *Overweight and obesity:* Body Mass Index (BMI) was calculated from self-reported height and weight and classified as not overweight, overweight, or obese using age- and sex-specific International Obesity Task Force thresholds.
2. *Longstanding physical health condition.* Cohort members reporting a longstanding condition affecting vision, hearing, mobility, dexterity or stamina were classified as having a longstanding physical health condition. Follow-up questions on its impact were processed in the same manner as for mental health conditions; the same limitation regarding non-specificity of impact applies.

3. *Physical activity.* Cohort members reported on how many days in the past week they had completed an hour of moderate-to-vigorous physical activity. Young people with no physical activity at all have been identified as being more likely to have mental health symptoms (32); therefore, cohort members were classified into 'no physical activity' (no days with an hour of physical activity in the past week; 1) and 'some physical activity' (at least one day with an hour of physical activity in the past week; 0).

*Health behaviours*. Health behaviours were based on self-reports at age 17.

1. *Addictive substances:* Following literature establishing weekly alcohol use in adolescence as being a predictor of problematic alcohol use in adulthood (33), cohort members' self-reports of their alcohol consumption were recoded to represent 'at least weekly alcohol use' ('6-9 times' in the past four weeks or more, which averages to at least once a week; 1) and 'not weekly alcohol use' (0). Repeated binge drinking was defined as consuming five or more drinks on three or more occasions in the past year (acknowledging that occasional instances might be considered developmentally normative and it is the repeated activity that is problematic; 34). Cohort members reporting smoking or vaping (or e-cigarettes) at least sometimes were classified as smoking and vaping, respectively. Cohort members also reported whether they had used cannabis in the past year and whether they had ever used hard drugs (e.g. cocaine, ecstasy, heroin, amphetamines or ketamine).
2. *Antisocial behaviours.* Cohort members reported whether they had engaged in any of several antisocial or criminal behaviours in the past year (e.g. theft, vandalism, fraud, cyberhacking, etc.). Any reported behaviour was coded as antisocial behaviour.

*Demographics.* Sex at birth, ethnic background, sexuality (any response other than heterosexual or mostly heterosexual was recoded as non-heterosexual) and nation of residence at age 17 were included. Four indicators measured socioeconomic background at age 14: highest parental occupation at home (professional/managerial versus any other response), highest parental post-18 qualification (a university degree or a diploma versus any other response), home ownership (yes/no), and whether household income was below 60% of the OECD-equivalised median (yes/no). Cohort members who reported their primary activity at age 17 as being in school or college, doing an apprenticeship or traineeship, training course or any kind of paid job were classified as not NEET at age 17. All others with non-missing responses were classified as NEET at age 17, in line with Office for National Statistics guidelines (20).

**Supplementary Material C: Prevalence rates of demographic factors and adolescent health exposures for the age 23 NEET and non-NEET status groups**

| | | | NEET | | Not NEET | |
|---|---|---|---|---|---|---|
| Category | Variable | Group / Level | n | Prevalence (%) | n | Prevalence (%) |
| Demographics | *Nation* | England | 693 | 86.2 | 4,757 | 83.2 |
| | | Wales | 140 | 4.8 | 915 | 4.8 |
| | | Scotland | 82 | 6.7 | 763 | 8.5 |
| | | N. Ireland | 71 | 2.3 | 714 | 3.4 |
| | *Sex* | Male | 481 | 53.9 | 3,261 | 49.7 |
| | | Female | 505 | 46.1 | 3,888 | 50.3 |
| | *Ethnicity* | Asian | 148 | 9.2 | 831 | 6.9 |
| | | Black | 42 | 4.1 | 235 | 2.5 |
| | | Mixed | 35 | 3.6 | 202 | 3.0 |
| | | Other | 7 | 0.4 | 35 | 0.4 |
| | | White | 753 | 82.7 | 5,844 | 87.1 |
| | *Sexuality* | Heterosexual | 793 | 84.2 | 6,293 | 90.2 |
| | | Non-heterosexual | 154 | 15.8 | 738 | 9.8 |
| | *Parents' highest qualification* | Post-18 qualification | 372 | 43.5 | 4,082 | 63.6 |
| | | No post-18 qualification | 496 | 56.5 | 2,587 | 36.4 |
| | *Parents' occupation* | Professional or managerial class | 271 | 49.2 | 3,402 | 59.0 |
| | | Not professional or managerial class | 362 | 50.8 | 2,623 | 41.0 |
| | *Family home ownership* | Owns home | 449 | 51.8 | 4,942 | 76.1 |
| | | Does not own home | 427 | 48.2 | 1,712 | 23.9 |
| | *Low income* | Yes | 430 | 41.1 | 1,498 | 15.9 |
| | | No | 466 | 58.9 | 5,249 | 84.1 |
| | *NEET at age 17* | Yes | 119 | 12.3 | 137 | 1.8 |
| | | No | 855 | 87.7 | 6,933 | 98.2 |
| Mental health difficulties | *Psychological distress* | Yes | 257 | 27.2 | 1,077 | 14.3 |
| | | No | 699 | 72.8 | 5,969 | 85.7 |
| | *Conduct problems* | Yes | 93 | 10.8 | 300 | 4.3 |
| | | No | 860 | 89.2 | 6,720 | 95.7 |
| | *Emotional symptoms* | Yes | 212 | 21.2 | 961 | 13.2 |
| | | No | 741 | 78.8 | 6,059 | 86.8 |
| | *Hyperactivity* | Yes | 210 | 22.5 | 928 | 13.2 |
| | | No | 743 | 77.5 | 6,092 | 86.8 |
| | *Self-harm in past 12 months* | Yes | 327 | 35.3 | 1,599 | 22.4 |
| | | No | 624 | 64.7 | 5,412 | 77.6 |
| | *Suicide attempt by age 17* | Yes | 137 | 14.6 | 456 | 6.4 |
| | | No | 809 | 85.4 | 6,553 | 93.6 |
| | | Yes | 135 | 16.8 | 364 | 5.6 |

| Category | Variable | Group / Level | NEET n | NEET Prevalence (%) | Not NEET n | Not NEET Prevalence (%) |
|---|---|---|---|---|---|---|
| | *Longstanding mental health condition…* | No | 839 | 83.2 | 6,704 | 94.4 |
| | *… which affects daily life* | Yes | 121 | 15.3 | 306 | 4.7 |
| | | No | 853 | 84.7 | 6,762 | 95.3 |
| Childhood neurodevelopmental conditions | *Autism* | Yes | 109 | 14.8 | 185 | 2.7 |
| | | No | 874 | 85.2 | 6,953 | 97.3 |
| | *ADHD* | Yes | 81 | 10.3 | 129 | 1.8 |
| | | No | 902 | 89.7 | 7,009 | 98.2 |
| Indicators of physical health | *Body mass index* | Normal | 566 | 63.9 | 4,766 | 71.4 |
| | | Overweight | 173 | 17.8 | 1,316 | 19.4 |
| | | Obesity | 151 | 18.3 | 678 | 9.2 |
| | *Longstanding physical health condition…* | Yes | 121 | 12.5 | 517 | 7.3 |
| | | No | 853 | 87.5 | 6,551 | 92.7 |
| | *… which affects daily life* | Yes | 92 | 10 | 314 | 4.4 |
| | | No | 882 | 90 | 6,753 | 95.6 |
| | *No physical activity* | Yes | 345 | 32.7 | 1,639 | 21.6 |
| | | No | 629 | 67.3 | 5,431 | 78.4 |
| Health behaviours | *Weekly alcohol use* | Yes | 58 | 7.4 | 871 | 14.1 |
| | | No | 894 | 92.6 | 6,158 | 85.9 |
| | *Repeated binge drinking* | Yes | 220 | 25.5 | 2,195 | 33.2 |
| | | No | 730 | 74.5 | 4,833 | 66.8 |
| | *Smoking* | Yes | 221 | 26.2 | 1,246 | 19.2 |
| | | No | 730 | 73.8 | 5,778 | 80.8 |
| | *Vaping* | Yes | 125 | 13.9 | 717 | 10.6 |
| | | No | 826 | 86.1 | 6,306 | 89.4 |
| | *Cannabis use in the past 12 months* | Yes | 268 | 32.8 | 1,924 | 29.8 |
| | | No | 685 | 67.2 | 5,106 | 70.2 |
| | *Hard drug use ever* | Yes | 87 | 12.1 | 566 | 9.3 |
| | | No | 869 | 87.9 | 6,472 | 90.7 |
| | *Antisocial behaviour in the past 12 months* | Yes | 136 | 15.6 | 706 | 10.8 |
| | | No | 820 | 84.4 | 6,340 | 89.2 |
| Mental health and neurodevelopmental difficulties | *Number of difficulties* | 0 | 482 | 45.8 | 4,771 | 67.6 |
| | | 1 | 214 | 23.3 | 1,408 | 19.4 |
| | | 2 | 136 | 14.5 | 633 | 8.4 |
| | | 3+ | 154 | 16.4 | 337 | 4.5 |
| | *Obesity and a difficulty* | Yes | 88 | 10.9 | 260 | 3.6 |
| | | No | 802 | 89.1 | 6,500 | 96.4 |

**Supplementary Material D:** *Relative risks and population attributable fractions of being NEET at age 23 (n = 8,135), adjusted for demographics, using non-imputed data.*

| Exposure | | Adjusted, weighted, non-imputed | | | |
|---|---|---|---|---|---|
| | | Relative risks | | Population attributable fractions | |
| | ***n*** | ***aRR*** | ***95% CI*** | ***PAF (%)*** | ***$p_c$*** |
| Mental health difficulties | | | | | |
| *Psychological distress* | 6,392 | 1.80* | 1.40, 2.33 | 12.12 | 27.19 |
| *Conduct problems* | 6,380 | 1.88* | 1.39, 2.54 | 5.06 | 10.8 |
| *Emotional symptoms* | 6,380 | 1.77* | 1.37, 2.28 | 9.21 | 21.24 |
| *Hyperactivity* | 6,380 | 1.51** | 1.19, 1.91 | 7.55 | 22.47 |
| *Self-harm in past 12 months* | 6,376 | 1.72* | 1.33, 2.21 | 14.74 | 35.27 |
| *Suicide attempt by age 17* | 6,371 | 1.97* | 1.48, 2.62 | 7.19 | 14.61 |
| *Longstanding mental health condition…* | 6,380 | 2.39* | 1.85, 3.09 | 9.78 | 16.79 |
| *… which affects daily life* | 6,380 | 2.52* | 1.93, 3.30 | 9.25 | 15.32 |
| Childhood neurodevelopmental conditions | | | | | |
| *Autism* | 6,395 | 3.60* | 2.69, 4.83 | 10.68 | 14.79 |
| *ADHD* | 6,395 | 3.25* | 2.38, 4.44 | 7.12 | 10.29 |
| Indicators of physical health | | | | | |
| *Overweight* | 6,143 | 1.00 | 0.77, 1.28 | n/a | 17.81 |
| *Obesity* | 6,143 | 1.54** | 1.18, 2.01 | 6.45 | 18.34 |
| *Longstanding physical health condition…* | 6,380 | 1.28 | 0.96, 1.71 | n/a | 12.55 |
| *… which affects daily life* | 6,379 | 1.61 | 1.16, 2.23 | n/a | 10 |
| *No physical activity* | 6,381 | 1.61* | 1.28, 2.03 | 12.43 | 32.69 |
| Health behaviours | | | | | |
| *Weekly alcohol use* | 6,377 | 0.60 | 0.42, 0.86 | n/a | 7.41 |
| *Repeated binge drinking* | 6,378 | 0.83 | 0.66, 1.06 | n/a | 25.53 |
| *Smoking* | 6,377 | 1.24 | 0.95, 1.60 | n/a | 26.18 |
| *Vaping* | 6,375 | 1.21 | 0.90, 1.61 | n/a | 13.85 |
| *Cannabis use in the past 12 months* | 6,380 | 1.17 | 0.93, 1.47 | n/a | 32.79 |
| *Hard drug use ever* | 6,381 | 1.50 | 1.02, 2.19 | n/a | 12.1 |
| *Antisocial behaviour in the past 12 months* | 6,395 | 1.52** | 1.18, 1.95 | 5.32 | 15.61 |
| Mental health and neurodevelopmental difficulties | | | | | |
| *1 difficulty* | 6,395 | 1.49 | 1.10, 2.03 | n/a | 23.32 |
| *2 difficulties* | 6,395 | 2.22* | 1.54, 3.19 | 7.97 | 14.52 |
| *3+ difficulties* | 6,395 | 3.55* | 2.54, 4.96 | 11.77 | 16.39 |
| *Obesity and a difficulty* | 6,143 | 2.01** | 1.38, 2.93 | 5.49 | 10.91 |
| NEET at age 17 | 6,381 | 4.19* | 3.17, 5.54 | 9.36 | 12.29 |

Note: Unweighted n's and prevalences are based on cases with valid data for each exposure. Weighted prevalence, relative risks and PAFs adjust for non-response to produce nationally representative estimates. Each relative risk was calculated independently. Adjusted relative risks controlled for nation, sex, ethnicity, sexuality and socioeconomic background. Reference categories are not reporting the exposure, except for overweight and obesity, where the reference is being not overweight. All p-values are corrected using the Holm procedure.

PAF estimates are not provided where the corresponding aRR was below 1 or if the association was non-significant after the Holm procedure.
Abbreviations: 95% CI = 95% confidence intervals; ADHD = attention deficit hyperactivity disorder; aRR = adjusted relative risks; PAF = population attributable fractions; $p_c$ = weighted prevalence of exposure among NEET cases.
Significance levels: * = $p < .001$; ** = $p < .05$.

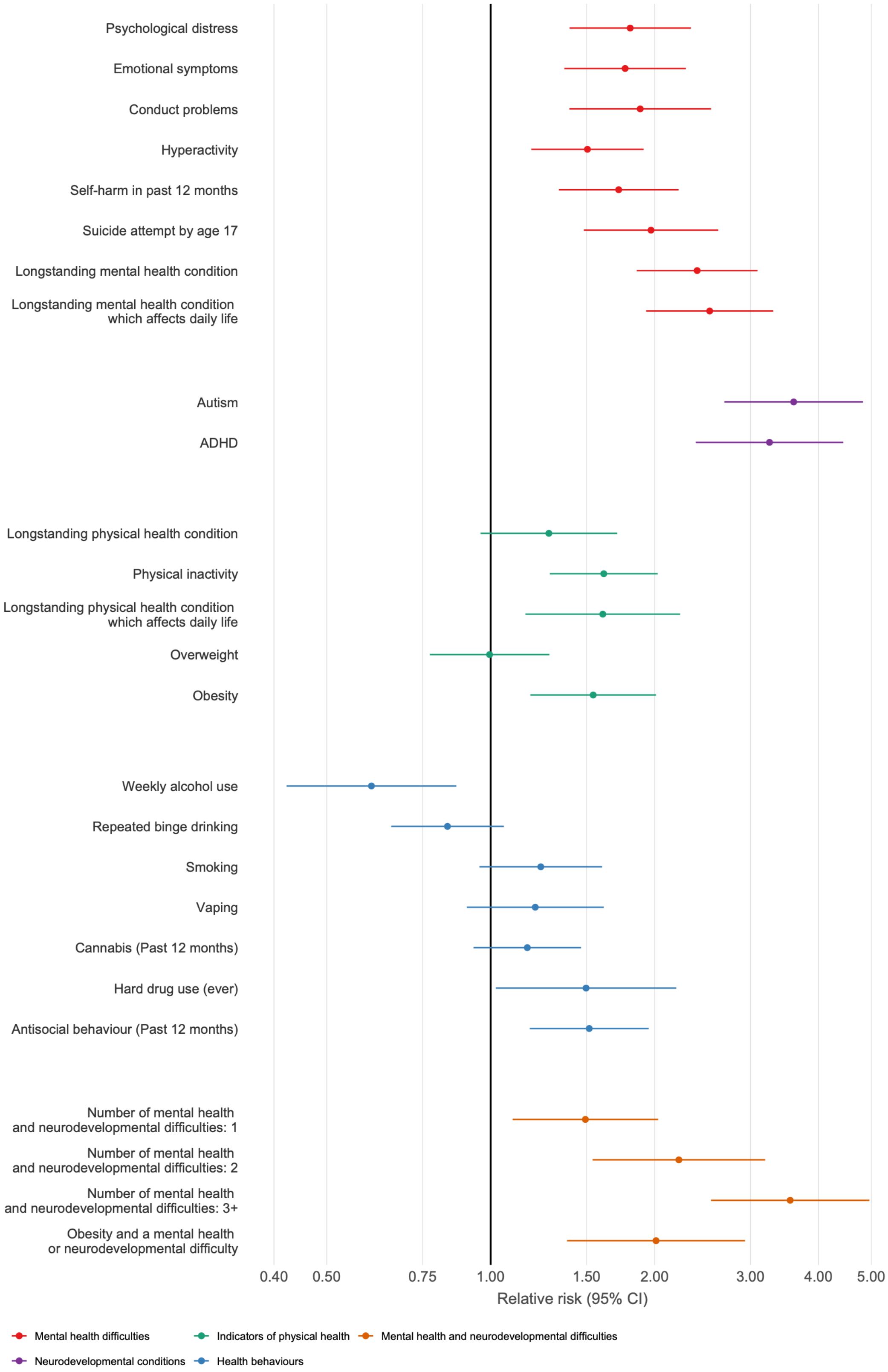

*Supplementary Figure 1.* Relative risks of being NEET at age 23 adjusting for demographics (nation, sex, ethnicity, sexuality, socioeconomic background). 95% CI = 95% confidence intervals. N = 8,135, non-imputed data.

**Supplementary Material E: Relative risks of health exposures for being NEET at age 23, unadjusted for demographic factors**

| Exposure | | *Unadjusted, weighted, non-imputed* | | *Unadjusted, weighted, imputed* | |
|---|---|---|---|---|---|
| | ***n*** | ***Relative risks*** | ***95% CI*** | ***Relative risks*** | ***95% CI*** |
| Mental health difficulties | | | | | |
| *Psychological distress* | 8,002 | 1.98* | 1.62, 2.43 | 1.97* | 1.62, 2.41 |
| *Conduct problems* | 7,973 | 2.28* | 1.86, 2.80 | 2.26* | 1.83, 2.81 |
| *Emotional symptoms* | 7,973 | 1.64* | 1.34, 2.01 | 1.60* | 1.30, 1.96 |
| *Hyperactivity* | 7,973 | 1.73* | 1.46, 2.06 | 1.73* | 1.45, 2.07 |
| *Self-harm in past 12 months* | 7,962 | 1.73* | 1.43, 2.09 | 1.72* | 1.43, 2.07 |
| *Suicide attempt by age 17* | 7,955 | 2.15* | 1.73, 2.66 | 2.14* | 1.73, 2.65 |
| *Longstanding mental health condition…* | 8,042 | 2.71* | 2.23, 3.29 | 2.67* | 2.20, 3.25 |
| *… which affects daily life* | 8,042 | 2.86* | 2.33, 3.51 | 2.82* | 2.29, 3.46 |
| Childhood neurodevelopmental conditions | | | | | |
| *Autism* | 8,121 | 3.96* | 3.22, 4.88 | 3.95* | 3.20, 4.86 |
| *ADHD* | 8,121 | 3.91* | 3.14, 4.87 | 3.89* | 3.12, 4.84 |
| Indicators of physical health | | | | | |
| *Overweight* | 7,650 | 1.02 | 0.82, 1.28 | 1.04 | 0.83, 1.30 |
| *Obesity* | 7,650 | 1.99* | 1.60, 2.46 | 2.02* | 1.64, 2.50 |
| *Longstanding physical health condition…* | 8,041 | 1.67* | 1.31, 2.13 | 1.66* | 1.31, 2.12 |
| *… which affects daily life* | 8,041 | 2.09* | 1.57, 2.78 | 2.07* | 1.56, 2.74 |
| *No physical activity* | 8,044 | 1.63* | 1.39, 1.92 | 1.63* | 1.39, 1.92 |
| Health behaviours | | | | | |
| *Weekly alcohol use* | 7,981 | 0.52* | 0.40, 0.69 | 0.52* | 0.39, 0.69 |
| *Repeated binge drinking* | 7,988 | 0.72** | 0.60, 0.86 | 0.71** | 0.59, 0.85 |
| *Smoking* | 7,975 | 1.41** | 1.18, 1.69 | 1.42** | 1.19, 1.69 |
| *Vaping* | 7,974 | 1.30 | 1.04, 1.64 | 1.31 | 1.05, 1.64 |
| *Cannabis use in the past 12 months* | 7,983 | 1.13 | 0.96, 1.33 | 1.13 | 0.96, 1.34 |
| *Hard drug use ever* | 7,994 | 1.30 | 0.96, 1.75 | 1.31 | 0.98, 1.76 |
| *Antisocial behaviour in the past 12 months* | 8,002 | 1.44** | 1.19, 1.74 | 1.44** | 1.19, 1.75 |
| Mental health and neurodevelopmental difficulties | | | | | |
| *1 difficulty* | 8,134 | 1.66* | 1.31, 2.10 | 1.70* | 1.35, 2.15 |
| *2 difficulties* | 8,134 | 2.24* | 1.69, 2.98 | 2.23* | 1.69, 2.95 |
| *3+ difficulties* | 8,134 | 3.90* | 3.04, 5.00 | 4.02* | 3.14, 5.15 |
| *Obesity and a difficulty* | 7,649 | 2.64* | 2.01, 3.49 | 2.75* | 2.10, 3.61 |
| NEET at age 17 | 8,044 | 4.38* | 3.64, 5.26 | 4.34* | 3.62, 5.22 |

Note: Weighted prevalence rates, relative risks and PAFs adjust for non-response to produce nationally representative estimates. Each relative risk was calculated independently. Reference categories are not reporting the condition or behaviour, except for overweight and obesity, where the reference is being not overweight. All p-values are corrected using the

Holm procedure. Abbreviations: 95% CI = 95% confidence interval; ADHD = attention deficit hyperactivity disorder. Significance levels: * = $p < .001$; ** = $p < .05$.

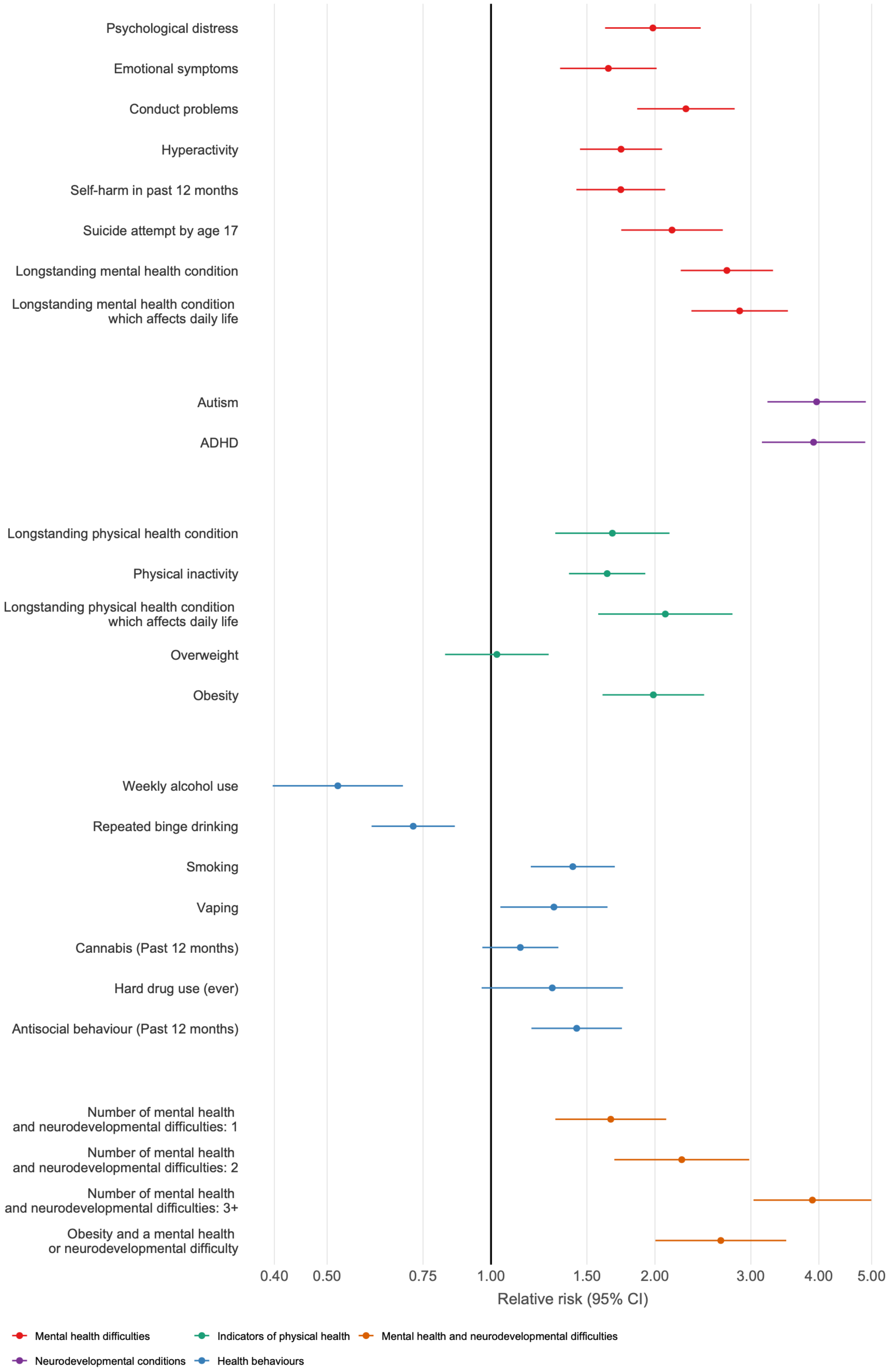

*Supplementary Figure 2.* Relative risks of being NEET at age 23 without adjusting for demographic factors, non-imputed data. 95% CI = 95% confidence intervals. N = 8,135.

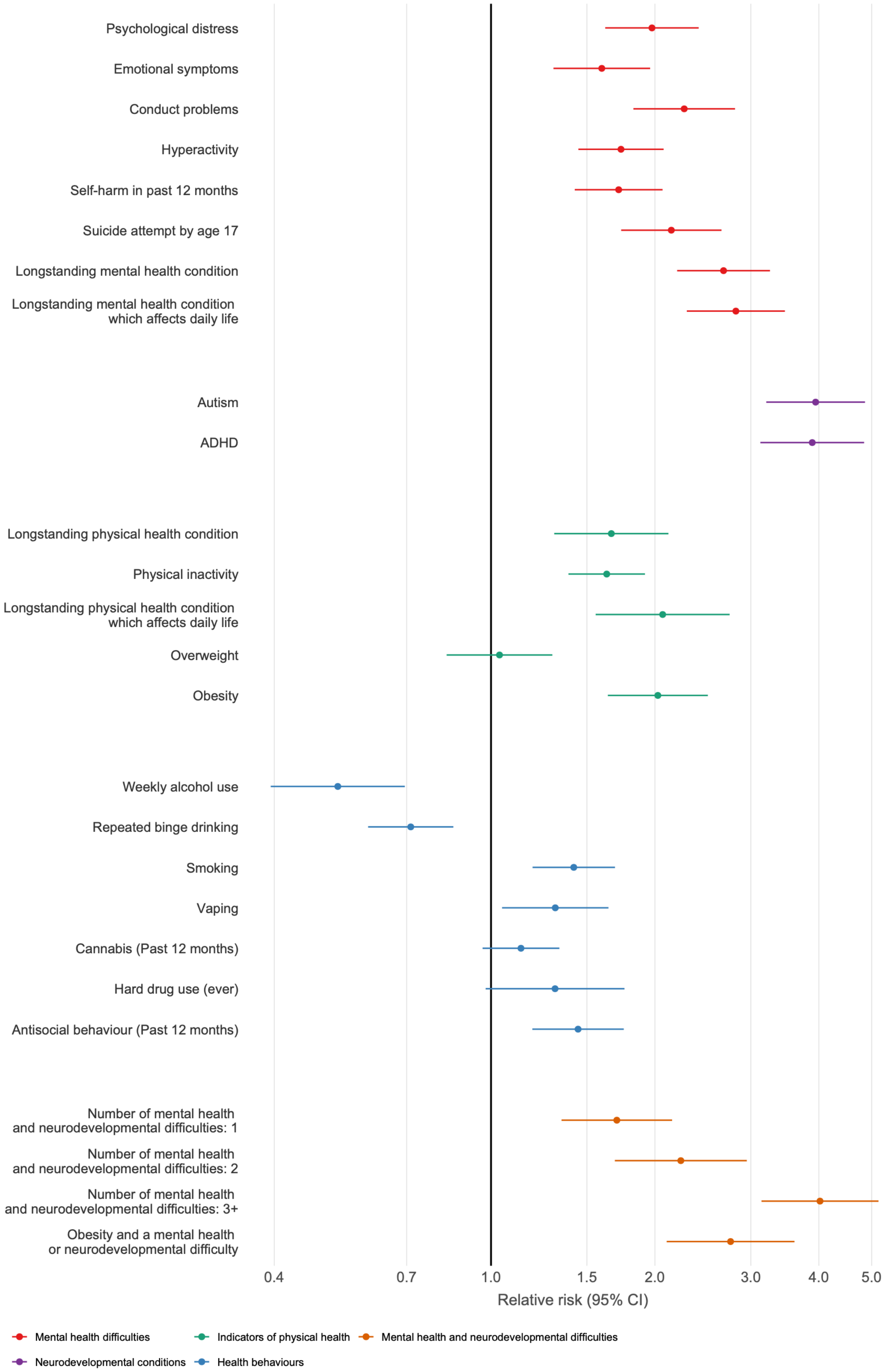

*Supplementary Figure 3.* Relative risks of being NEET at age 23 without adjusting for demographic factors, imputed data. 95% CI = 95% confidence intervals. N = 8,135.

**Supplementary Material F: Sensitivity Analyses**

*Supplementary Table F1* shows the magnitude of the relative risks among cohort members who were not identified as NEET at age 17 (the majority of the sample, approximately 97%, which were similar to those in the full sample and did not show a consistent pattern of differences.

*Supplementary Table F2* shows that most aRRs and PAFs were larger for economic inactivity than unemployment, suggesting that health exposures at age 17 were more strongly linked to being inactive than unemployed. Autism was the main exception and had a comparatively stronger association with unemployment than economic inactivity (aRRs = 3.87 vs 3.24). The largest PAFs for being economically inactive were for self-harm in the past 12 months (28.89%), having a longstanding mental health condition (26.63%) and psychological distress (25.11%). This suggests that poor health may be more strongly linked to disengagement from the labour market than to difficulties finding employment. However, findings should be interpreted cautiously, as some cohort members may have misclassified their employment status, given that the term 'unemployed' is not always used colloquially to mean actively seeking work.

*Supplementary Table F1: Relative risks of being NEET at age 23 presented with and without cohort members who were already identified as NEET at age 17.*

| **Exposure** | **Adjusted and weighted** | | | | |
|---|---|---|---|---|---|
| | **Relative risks without NEET at age 17 (non-imputed)** | | **Relative risks without NEET at age 17 (imputed)** | | **Difference from analytic sample imputed aRR** |
| | ***aRR*** | ***95% CI*** | ***aRR*** | ***95% CI*** | **aRR** |
| Mental health difficulties | | | | | |
| *Psychological distress* | 1.75* | 1.34, 2.29 | 1.72* | 1.37, 2.16 | -0.06 |
| *Conduct problems* | 1.78** | 1.26, 2.51 | 1.75* | 1.38, 2.22 | +0.01 |
| *Emotional symptoms* | 1.74* | 1.33, 2.27 | 1.59** | 1.26, 2.00 | 0 |
| *Hyperactivity* | 1.44** | 1.14, 1.83 | 1.52* | 1.27, 1.81 | 0 |
| *Self-harm in past 12 months* | 1.68* | 1.31, 2.17 | 1.56* | 1.28, 1.91 | -0.03 |
| *Suicide attempt by age 17* | 1.87* | 1.36, 2.56 | 1.56** | 1.21, 2.02 | -0.05 |
| *Longstanding mental health condition…* | 2.27* | 1.69, 3.05 | 2.12* | 1.67, 2.70 | -0.03 |
| *… which affects daily life* | 2.34* | 1.70, 3.21 | 2.17* | 1.67, 2.82 | -0.06 |
| <u>Childhood neurodevelopmental conditions</u> | | | | | |
| *Autism* | 3.49* | 2.56, 4.76 | 3.01* | 2.34, 3.86 | +0.07 |
| *ADHD* | 2.93* | 1.97, 4.35 | 2.71* | 2.04, 3.60 | -0.03 |
| <u>Indicators of physical health</u> | | | | | |
| *Overweight* | 1.03 | 0.79, 1.35 | 0.94 | 0.74, 1.20 | 0 |
| *Obesity* | 1.48** | 1.11, 1.97 | 1.50** | 1.18, 1.89 | +0.02 |
| *Longstanding physical health condition…* | 1.13 | 0.81, 1.56 | 1.43 | 1.08, 1.90 | -0.05 |
| *… which affects daily life* | 1.31 | 0.88, 1.95 | 1.69 | 1.18, 2.42 | -0.07 |
| *No physical activity* | 1.41** | 1.09, 1.82 | 1.27 | 1.05, 1.55 | -0.14 |
| <u>Health behaviours</u> | | | | | |
| *Weekly alcohol use* | 0.59** | 0.40, 0.86 | 0.62 | 0.45, 0.86 | -0.02 |
| *Repeated binge drinking* | 0.81 | 0.63, 1.04 | 0.83 | 0.68, 1.02 | -0.01 |
| *Smoking* | 1.13 | 0.86, 1.48 | 1.20 | 0.99, 1.44 | -.010 |
| *Vaping* | 1.15 | 0.84, 1.58 | 1.09 | 0.86, 1.39 | -0.07 |
| *Cannabis use in the past 12 months* | 1.19 | 0.96, 1.49 | 1.13 | 0.96, 1.33 | 0 |
| *Hard drug use ever* | 1.38 | 0.97, 1.95 | 1.30 | 0.97, 1.75 | -0.04 |
| *Antisocial behaviour in the past 12 months* | 1.36 | 1.01, 1.83 | 1.29 | 1.03, 1.60 | -0.09 |
| <u>Mental health and neurodevelopmental difficulties</u> | | | | | |
| *1 difficulty* | 1.53** | 1.14, 2.07 | 1.67* | 1.31, 2.13 | -0.06 |
| *2 difficulties* | 2.21* | 1.53, 3.19 | 2.04* | 1.51, 2.75 | 0 |
| *3+ difficulties* | 3.27* | 2.30, 4.63 | 3.08* | 2.29, 4.14 | -0.08 |
| *Obesity and a difficulty* | 1.81** | 1.24, 2.64 | 1.98* | 1.45, 2.71 | -0.03 |

Note: Weighted relative risks adjust for non-response to produce nationally representative estimates. Each relative risk was calculated independently. Adjusted relative risks controlled for nation, sex, ethnicity, sexuality and socioeconomic background. Reference categories are not reporting the condition or behaviour, except for overweight and obesity, where the reference is being not overweight, including being underweight. All p-values are corrected using the Holm procedure. Differences were calculated by subtracting the full analytic sample imputed aRR from the imputed aRR after excluding cohort members who were NEET at age 17.

Abbreviations: 95% CI = 95% confidence intervals; ADHD = attention deficit hyperactivity disorder; aRR = adjusted relative risks. Significance levels: * = $p < .001$; ** = $p < .05$.

*Supplementary Table F2:* Relative risks and population attributable fractions calculated separately for being unemployed and economically inactive at age 23, with non-imputed data.

| **Exposure** | **Adjusted relative risks** | | | | **Population attributable fractions** | | | |
|---|---|---|---|---|---|---|---|---|
| | **Unemployed** | | **Economically inactive** | | **Unemployed** | | **Economically inactive** | |
| | ***aRR*** | ***95% CI*** | ***aRR*** | ***95% CI*** | ***PAF (%)*** | $p_c$ | ***PAF (%)*** | $p_c$ |
| Mental health difficulties | | | | | | | | |
| *Psychological distress* | 1.42 | 1.03, 1.96 | 2.75* | 1.73, 4.36 | n/a | 20.59 | 25.11 | 39.48 |
| *Conduct problems* | 1.40 | 0.93, 2.08 | 2.99* | 1.8, 4.98 | n/a | 8.65 | 9.98 | 14.99 |
| *Emotional symptoms* | 1.45 | 1.06, 1.98 | 2.43** | 1.57, 3.75 | n/a | 16.46 | 18.01 | 30.60 |
| *Hyperactivity* | 1.15 | 0.85, 1.55 | 2.47* | 1.65, 3.69 | n/a | 18.69 | 17.75 | 29.87 |
| *Self-harm in past 12 months* | 1.37 | 1.06, 1.76 | 2.73** | 1.58, 4.71 | n/a | 29.76 | 28.89 | 45.62 |
| *Suicide attempt by age 17* | 1.17 | 0.76, 1.82 | 3.52* | 2.35, 5.29 | n/a | 8.89 | 18.22 | 25.45 |
| *Longstanding mental health condition…* | 1.33 | 0.91, 1.95 | 4.65* | 3.02, 7.16 | n/a | 7.68 | 26.63 | 33.93 |
| *… which affects daily life* | 1.42 | 0.95, 2.12 | 4.77* | 3.05, 7.47 | n/a | 6.81 | 24.75 | 31.32 |
| <u>Childhood neurodevelopmental conditions</u> | | | | | | | | |
| *Autism* | 3.87* | 2.63, 5.72 | 3.24** | 1.85, 5.69 | 10.53 | 14.19 | 11.03 | 15.95 |
| *ADHD* | 2.62** | 1.59, 4.30 | 4.69* | 2.80, 7.84 | 4.96 | 8.03 | 11.53 | 14.66 |
| <u>Indicators of physical health</u> | | | | | | | | |
| *Overweight* | 0.95 | 0.68, 1.33 | 1.11 | 0.69, 1.78 | n/a | 16.25 | n/a | 22.10 |
| *Obesity* | 1.40 | 1.02, 1.93 | 1.85 | 1.13, 3.05 | n/a | 16.43 | n/a | 22.37 |
| *Longstanding physical health condition…* | 0.80 | 0.50, 1.29 | 2.36** | 1.51, 3.69 | n/a | 8.14 | 12.00 | 20.83 |
| *… which affects daily life* | 0.87 | 0.52, 1.47 | 3.13* | 1.87, 5.23 | n/a | 5.70 | 12.31 | 18.10 |
| *No physical activity* | 1.45 | 1.11, 1.9 | 2.01** | 1.41, 2.88 | n/a | 30.22 | 18.79 | 37.32 |
| <u>Health behaviours</u> | | | | | | | | |
| *Weekly alcohol use* | 0.65 | 0.42, 1.00 | 0.49 | 0.22, 1.09 | n/a | 8.92 | n/a | 4.60 |
| *Repeated binge drinking* | 0.83 | 0.62, 1.12 | 0.84 | 0.56, 1.25 | n/a | 25.43 | n/a | 25.73 |
| *Smoking* | 1.02 | 0.75, 1.37 | 1.75 | 1.05, 2.94 | n/a | 22.61 | n/a | 32.81 |

| | | | | | | | | |
|---|---|---|---|---|---|---|---|---|
| *Vaping* | 1.08 | 0.76, 1.52 | 1.49 | 0.86, 2.58 | n/a | 13.41 | n/a | 14.67 |
| *Cannabis use in the past 12 months* | 1.1 0 | 0.87, 1.40 | 1.31 | 0.81, 2.13 | n/a | 30.98 | n/a | 36.17 |
| *Hard drug use ever* | 1.27 | 0.88, 1.82 | 2.07 | 0.96, 4.47 | n/a | 10.61 | n/a | 14.90 |
| *Antisocial behaviour in the past 12 months* | 1.27 | 0.90, 1.80 | 2.14** | 1.42, 3.22 | n/a | 15.06 | 8.60 | 16.65 |
| <u>Mental health and neurodevelopmental difficulties</u> | | | | | | | | |
| *1 difficulty* | 1.43 | 1.07, 1.92 | 1.79 | 0.81, 3.93 | n/a | 23.81 | n/a | 22.41 |
| *2 difficulties* | 1.95** | 1.34, 2.87 | 3.25** | 1.51, 7.02 | 6.20 | 12.69 | 12.42 | 17.93 |
| *3+ difficulties* | 2.29** | 1.43, 3.64 | 7.13* | 3.56, 14.27 | 6.48 | 11.50 | 21.98 | 25.57 |
| *Obesity and a difficulty* | 1.85 | 1.24, 2.77 | 2.19 | 1.10, 4.36 | n/a | 9.98 | 6.84 | 12.60 |
| <u>NEET at age 17</u> | 3.23* | 2.13, 4.90 | 5.75* | 3.16, 10.49 | 7.49 | 10.85 | 12.42 | 14.99 |

Note: Weighted prevalence rates, relative risks and PAFs adjust for non-response to produce nationally representative estimates. Each relative risk was calculated independently. Adjusted relative risks controlled for nation, sex, ethnicity, sexuality and socioeconomic background. Reference categories are not reporting the condition or behaviour, except for overweight and obesity, where the reference is being not overweight, including being underweight. All p-values are corrected using the Holm procedure. PAF estimates are not provided where the corresponding aRR was below 1 or if the association was non-significant after the Holm procedure.
Abbreviations: 95% CI = 95% confidence intervals; ADHD = attention deficit hyperactivity disorder; aRR = adjusted relative risks; PAF = population attributable fractions; $p_c$ = weighted prevalence of exposure among NEET cases. Significance levels: * = $p < .001$; ** = $p < .05$.

**Supplementary Material G. Weighted, exposure-specific adjusted relative risks and population attributable fractions of being NEET at age 23 (n = 8,135), divided by sex, with accompanying tests of interaction effects, non-imputed**

| Exposure | Adjusted relative risks | | | | | Population attributable fractions | | | |
|---|---|---|---|---|---|---|---|---|---|
| | Female | | Male | | Interaction | Female | | Male | |
| | *aRR* | *95% CI* | *aRR* | *95% CI* | *p-value* | *PAF (%)* | $p_c$ | *PAF (%)* | $p_c$ |
| Mental health difficulties | | | | | | | | | |
| *Psychological distress* | 1.99* | 1.45, 2.73 | 1.52 | 1.02, 2.26 | .098 | 19.41 | 38.97 | n/a | 16.93 |
| *Conduct problems* | 2.05** | 1.37, 3.06 | 1.77 | 1.19, 2.65 | .532 | 5.03 | 9.82 | n/a | 11.67 |
| *Emotional symptoms* | 1.79** | 1.34, 2.38 | 1.65 | 1.12, 2.48 | .446 | 14.63 | 33.25 | n/a | 10.57 |
| ***Hyperactivity*** | **1.97*** | **1.51, 2.56** | **1.14** | **0.79, 1.65** | **.004** | **11.87** | **24.14** | **n/a** | **20.99** |
| *Self-harm in past 12 months* | 1.81** | 1.26, 2.59 | 1.62 | 1.18, 2.23 | .294 | 19.76 | 44.22 | n/a | 27.51 |
| *Suicide attempt by age 17* | 2.13** | 1.47, 3.08 | 1.55 | 0.97, 2.48 | .117 | 10.92 | 20.60 | n/a | 9.48 |
| *Longstanding mental health condition…* | 2.20* | 1.59, 3.05 | 2.70* | 1.72, 4.24 | .767 | 11.58 | 21.20 | 8.21 | 13.03 |
| *… which affects daily life* | 2.29* | 1.62, 3.24 | 2.91* | 1.83, 4.64 | .725 | 10.60 | 18.85 | 8.08 | 12.31 |
| Childhood neurodevelopmental conditions | | | | | | | | | |
| *Autism* | 3.11* | 1.83, 5.28 | 4.11* | 2.95, 5.71 | .755 | 4.65 | 6.85 | 16.27 | 21.51 |
| *ADHD* | 2.7** | 1.44, 5.04 | 3.46* | 2.41, 4.95 | .977 | 2.88 | 4.58 | 10.75 | 15.12 |
| Indicators of physical health | | | | | | | | | |
| *Overweight* | 0.97 | 0.71, 1.31 | 1.02 | 0.70, 1.49 | .929 | n/a | 18.07 | n/a | 17.61 |
| *Obesity* | 1.35 | 0.90, 2.02 | 1.71 | 1.20, 2.43 | .929 | n/a | 18.69 | n/a | 18.05 |
| *Longstanding physical health condition…* | 1.33 | 0.93, 1.91 | 1.21 | 0.66, 2.21 | .672 | n/a | 13.87 | n/a | 11.42 |
| *… which affects daily life* | 1.48 | 0.98, 2.23 | 1.75 | 0.90, 3.41 | .842 | n/a | 11.64 | n/a | 8.60 |
| *No physical activity* | 1.40 | 1.02, 1.91 | 1.91** | 1.39, 2.62 | .340 | n/a | 36.61 | 13.95 | 29.34 |
| Health behaviours | | | | | | | | | |
| *Weekly alcohol use* | 0.73 | 0.45, 1.18 | 0.53 | 0.31, 0.91 | .513 | n/a | 6.88 | n/a | 7.87 |
| *Repeated binge drinking* | 0.86 | 0.65, 1.14 | 0.80 | 0.55, 1.17 | .866 | n/a | 25.95 | n/a | 25.17 |
| *Smoking* | 1.45 | 1.09, 1.93 | 1.04 | 0.70, 1.55 | .092 | n/a | 30.4 | n/a | 22.50 |
| *Vaping* | 1.37 | 0.88, 2.13 | 1.08 | 0.75, 1.57 | .371 | n/a | 12.10 | n/a | 15.38 |
| *Cannabis use in the past 12 months* | 1.15 | 0.87, 1.53 | 1.15 | 0.83, 1.59 | .893 | n/a | 31.99 | n/a | 33.48 |
| *Hard drug use ever* | 1.63 | 1.14, 2.32 | 1.4 0 | 0.84, 2.32 | .465 | n/a | 10.32 | n/a | 13.65 |

| | | | | | | | | | |
|---|---|---|---|---|---|---|---|---|---|
| *Antisocial behaviour in the past 12 months* | 1.53 | 1.04, 2.25 | 1.50 | 1.05, 2.15 | .922 | n/a | 13.13 | n/a | 17.77 |
| Mental health and neurodevelopmental difficulties | | | | | | | | | |
| *1 difficulty* | 1.10 | 0.71, 1.70 | 1.90** | 1.37, 2.65 | .192 | n/a | 22.80 | 11.26 | 23.76 |
| *2 difficulties* | 2.10** | 1.31, 3.35 | 2.29** | 1.39, 3.76 | .192 | 8.61 | 16.44 | 7.24 | 12.87 |
| *3+ difficulties* | 3.41* | 2.19, 5.31 | 3.29* | 2.11, 5.15 | .192 | 14 | 19.81 | 9.38 | 13.47 |
| *Obesity and a difficulty* | 1.54 | 0.94, 2.53 | 2.44** | 1.54, 3.88 | .192 | n/a | 12.33 | 5.77 | 9.77 |
| NEET at age 17 | 4.35* | 2.92, 6.49 | 3.89* | 2.46, 6.15 | .442 | 9.72 | 12.62 | 8.91 | 12.00 |

Note: Weighted prevalence rates, relative risks and PAFs adjust for non-response to produce nationally representative estimates. Each relative risk was calculated independently. Adjusted relative risks controlled for nation, sex, ethnicity, sexuality and socioeconomic background. Reference categories are not reporting the condition or behaviour, except for overweight and obesity, where the reference is being not overweight, including being underweight. Rows highlighted in bold signify a significant interaction effect of sex. All p-values are corrected using the Holm procedure, except for interaction effects. PAF estimates are not provided where the corresponding aRR was below 1 or if the association was non-significant after the Holm procedure.
Abbreviations: 95% CI = 95% confidence interval; ADHD = attention deficit hyperactivity disorder; aRR = adjusted relative risks; PAF = population attributable fractions; $p_c$ = weighted prevalence of exposure among NEET cases. Significance levels: * = $p < .001$; ** = $p < .05$.
